# Optoacoustic molecular signatures of muscle involvement in facioscapulohumeral dystrophy compared to conventional imaging

**DOI:** 10.64898/2026.09.04.26362282

**Authors:** Mauro Monforte, Sara Bortolani, Davide Marchese, Beatrice Ravera, Carmine Di Marco, Eleonora Torchia, Neal C. Burton, Thomas Sardella, Tommaso Pirronti, Tommaso Tartaglione, Marco De Spirito, Enzo Ricci, Giorgio Tasca

## Abstract

**Background and Objectives:** Facioscapulohumeral muscular dystrophy (FSHD) is characterised by asynchronous muscle degeneration and marked pathological heterogeneity. While conventional MRI can detect fatty replacement and oedema-like abnormalities, it provides limited insight into other relevant biological processes such as extracellular matrix remodelling and tissue perfusion. This study investigated whether multispectral optoacoustic tomography (MSOT), an emerging imaging technique that non-invasively interrogates tissue composition through endogenous optical absorbers, can provide complementary molecular information on muscle involvement in FSHD.

**Methods:** MSOT was performed on 568 muscles from 15 genetically confirmed FSHD patients and 9 healthy volunteers using a standardized protocol. The analysis included single-wavelength signal intensity and spectrally unmixed signals related to lipid, collagen, deoxygenated haemoglobin, oxygenated haemoglobin, total haemoglobin, and tissue oxygen saturation. Patients also underwent conventional muscle MRI together with clinical strength and disease severity assessments. Feasibility, reproducibility, discrimination between patients and controls, and associations with MRI and clinical measures were evaluated.

**Results:** MSOT acquisition was well tolerated and highly reproducible (intra-and inter-rater ICC = 0.88). FSHD muscles displayed a distinct optoacoustic profile characterised by reduced signal intensity at wavelengths below 900 nm, increased lipid signal and reduced oxygenated and total haemoglobin signals. Lipid signal discriminated FSHD from control muscles (AUC 0.830, p = 0.008) and increased with MRI-defined fatty replacement (p < 0.0001). Short tau inversion recovery (STIR)-positive muscles on MRI exhibited a distinct molecular phenotype, with higher collagen-associated and lipid signals together with increased deoxygenated and total haemoglobin signals, consistent with active tissue remodelling. Although FSHD muscles classified as normal by conventional imaging showed group-level optoacoustic profiles comparable to controls, 13 of 141 (9%) demonstrated at least one abnormal MSOT-derived parameter. Reduced haemoglobin-related signals correlated with greater global disease severity, whereas increased lipid signal was associated with lower muscle strength.

**Discussion:** MSOT is a feasible and reproducible imaging technique in FSHD that provides complementary molecular information beyond conventional structural imaging. Its ability to detect signatures related to fatty replacement, STIR positivity, functional impairment, and isolated abnormalities in radiologically normal muscles supports further longitudinal studies of MSOT-derived readouts as potential biomarkers of disease activity, progression, and therapeutic response.

## Introduction

Facioscapulohumeral muscular dystrophy (FSHD) is one of the most common inherited myopathies in adults and results from inappropriate skeletal muscle expression of the DUX4 retrogene^1–3^. Despite a common molecular cause, disease expression is remarkably heterogeneous^4–7^. Individual muscles undergo degeneration in an asynchronous manner, with active disease, chronic fibro-fatty replacement and relatively preserved muscle frequently coexisting within the same patient^8^. This spatial and temporal heterogeneity represents a major challenge for disease monitoring and for the development of sensitive biomarkers capable of capturing biologically meaningful changes over time.

Muscle magnetic resonance imaging (MRI) has transformed the evaluation of FSHD in both clinical practice and research. T1-weighted sequences provide a robust assessment of fatty replacement, whereas short tau inversion recovery (STIR) sequences identify oedema-like signal abnormalities associated with disease activity^9–11^. MRI has therefore become an essential tool for patient stratification, natural history studies and clinical trials^12,13^. Nevertheless, conventional MRI primarily reflects structural changes and provides limited direct information on other pathological processes that contribute to muscle degeneration.

Increasing evidence indicates that extracellular matrix remodelling, fibrosis and alterations in muscle perfusion are integral components of FSHD pathophysiology^14,15^. Histopathological studies have demonstrated progressive collagen deposition within affected muscles^16^, while MRI-guided biopsy studies have linked STIR-positive muscles with inflammatory changes, activation of DUX4 target genes and pro-angiogenic changes^17,18^. Likewise, abnormalities in muscle oxygenation and vascular physiology have been described in FSHD, suggesting that changes in tissue perfusion may accompany disease progression^19,20^. Conventional structural imaging, including MRI and ultrasound-based techniques, primarily visualize architectural or anatomical alterations, lacking the ability to provide specific molecular-level details about these processes. Ultrasound can detect alterations in muscle echogenicity but lacks specificity, as increased echogenicity may reflect fat infiltration, fibrosis, oedema or combinations of these processes^21–23^.

Multispectral optoacoustic tomography (MSOT) is an emerging imaging technique that combines optical excitation with ultrasound detection to generate real-time maps of endogenous tissue chromophores^24^. By exploiting wavelength-dependent optical absorption, MSOT enables non-invasive estimation of tissue components including lipids, collagen-associated signals and haemoglobin-related parameters, thereby providing information on tissue composition and oxygenation without exogenous contrast agents. Unlike conventional imaging, MSOT therefore has the potential to interrogate molecular and functional aspects of muscle pathology in vivo^25^.

Initial studies have demonstrated the feasibility of MSOT in neuromuscular disorders, including Duchenne muscular dystrophy, spinal muscular atrophy and Pompe disease, where the technique identified disease-associated alterations in lipid and collagen-associated signals^26–28^. Beyond neuromuscular disease, MSOT has also shown promise for assessing tissue perfusion in vascular^29,30^ and immune-mediated disorders. Changes in MSOT haemoglobin-related parameters corresponded to inflammatory activity in patients with Crohn’s disease^31^ and arthritis^32,33^. Whether these molecular readouts provide clinically relevant information in FSHD, however, remains unknown.

In this pilot study, we investigated the feasibility and reproducibility of MSOT in FSHD and examined its relationship with conventional imaging and clinical measures. Specifically, we asked whether MSOT identifies molecular signatures associated with different stages of muscle involvement, whether these signatures complement established MRI biomarkers and reflected ultrasound computed tomography (RUCT)-derived morphology, and whether optoacoustic abnormalities can be detected in muscles that appear normal on conventional imaging.

## Materials and methods

### Study population

This prospective observational study was conducted at the Fondazione Policlinico Universitario A. Gemelli IRCCS, Rome, Italy. Eligible participants were adults (18–65 years) with genetically confirmed FSHD1 or FSHD2 who had undergone routine upper and lower-limb muscle MRI within 60 days before enrolment. Healthy volunteers without neuromuscular disease were recruited as controls. Exclusion criteria included pregnancy and medical conditions potentially affecting skeletal muscle composition, including peripheral neuropathy, radiculopathy, uncontrolled diabetes mellitus, and acute or chronic systemic inflammatory disorders. Individual muscle measurements were not performed when local factors (e.g. tattoos, scars, skin lesions or recent trauma) could interfere with optoacoustic acquisition. Demographic and anthropometric data were collected for all participants. Clinical assessment included neurological examination, hand-held dynamometry (Hoggan microFET®) and global disease severity using the FSHD Clinical Severity Scale (CSS)^34^. Genetic data included EcoRI fragment length for FSHD1 and D4Z4 methylation status together with *SMCHD1* variants for FSHD2.

### MSOT acquisition

MSOT examinations were performed using a CE-marked hybrid reflected ultrasound computed tomography (RUCT)/MSOT system (MSOT Acuity Echo, iThera Medical GmbH, Munich, Germany; Figure 1A). Participants rested supine for at least 30 minutes before imaging to minimise potential effects of recent muscle activity. Images were acquired at twelve wavelengths (700, 730, 760, 800, 850, 910, 930, 950, 980, 1030, 1080 and 1100 nm). A selective frame-averaging algorithm averaged seven consecutive wavelength cycles. Imaging was performed using a handheld 4-MHz probe (256 detector elements; 40 × 40 mm field of view; approximately 150 μm spatial resolution), positioned perpendicular to the skin with ultrasound gel as coupling medium. Co-registered RUCT images provided anatomical guidance during acquisition. The following muscles were examined bilaterally: trapezius, deltoid, biceps brachii, upper and lower rectus abdominis, rectus femoris, proximal and distal vastus lateralis, biceps femoris, tibialis anterior, medial gastrocnemius and lumbar paraspinal muscles. Each muscle was scanned twice during the same examination according to predefined anatomical landmarks (Supplementary document). Examination tolerability was assessed by recording adverse events during and immediately after imaging.

**FIGURE 1.**
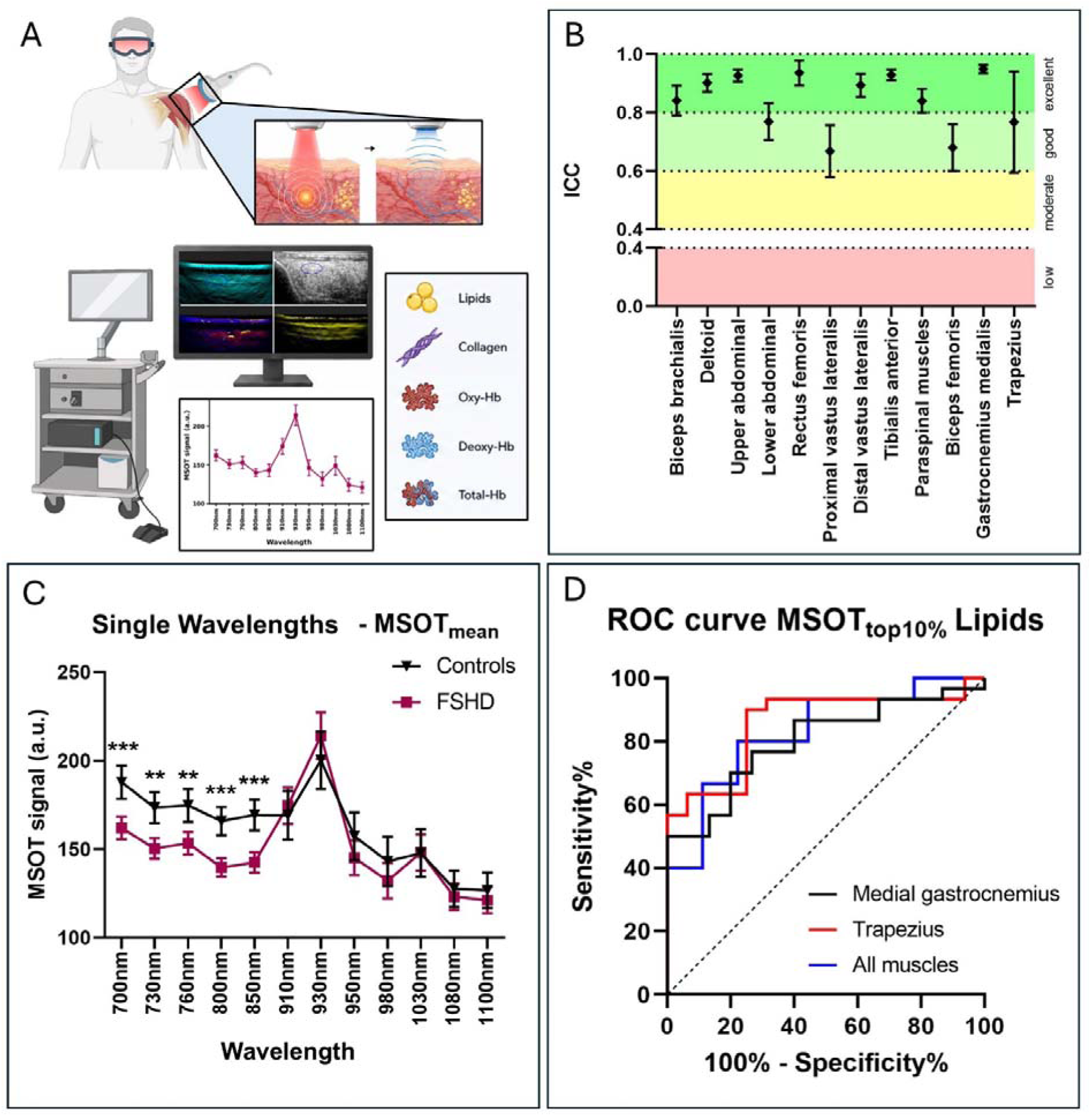
MSOT showed excellent reproducibility and identified a characteristic optoacoustic profile in FSHD muscle. (**A**) Schematic representation of reflected ultrasound computed tomography and multispectral optoacoustic tomography acquisition and spectral unmixing. (**B**) Repeat-acquisition reproducibility across individual muscles. Intraclass correlation coefficient (ICC) values for repeated MSOT acquisitions by muscle. Shaded bands indicate conventional ICC interpretation ranges. Repeat-acquisition reproducibility was excellent across all muscles, with ICC values consistently within the good-to-excellent range. The highest reproducibility was observed for the rectus femoris and medial gastrocnemius, whereas the proximal vastus lateralis and biceps femoris showed the lowest ICC values while remaining within the good reproducibility range. (**C**) Pooled MSOT_mean_ single-wavelength signal intensity in control and FSHD muscles. Patient muscles showed reduced signal at shorter wavelengths and a relative increase around 930 nm. (**D**) Receiver operating characteristic curves for MSOT_top10%_ lipid signal in all muscles and in selected clinically relevant muscles. Error bars indicate 95% C.I. * p < 0.05, ** p < 0.01, *** p < 0.001, **** p < 0.0001. Panel A was partially created with the use of BioRender.com and ChatGPT, GPT 5.5, OpenAI.

### MSOT image analysis

MSOT datasets were analysed offline using cLabs software (version 2.65, iThera Medical GmbH). A trained investigator (DM), blinded to clinical information, manually placed an elliptical region of interest (ROI) immediately beneath the muscle fascia on co-registered RUCT images. ROIs included only muscle tissue while excluding fascia, bone, tendons and large vessels. ROI size ranged from 20 to 40 mm². RUCT images were graded according to the Heckmatt scale (grades 1– 4)^23^, with grades 2 and 3 analysed together because of limited numbers.

For each ROI, both single-wavelength (SWL) signal intensity and spectrally unmixed signals were extracted. Spectral unmixing generated collagen-associated signal, lipid signal, deoxygenated haemoglobin (Hb), oxygenated haemoglobin (HbO□), total haemoglobin (HbT) and tissue oxygen saturation (mSO□). Haemoglobin parameters were calculated using wavelengths between 700 and 850 nm, whereas collagen-associated and lipid signals used the complete spectral range. Results are reported as arbitrary units (a.u.). Two quantitative metrics were analysed for each parameter: the mean ROI signal (MSOT_mean_) and the mean value of the highest 10% of pixels within the ROI (MSOT _top10%_).

### MRI acquisition and analysis

Muscle MRI was performed using a 1.5-T scanner (Magnetom Espree, Siemens) following previously published acquisition protocols^10,35^. Axial T1-weighted sequences were used to assess fatty replacement, while fat-suppressed STIR sequences evaluated oedema-like signal abnormalities. Images were independently reviewed by two experienced raters (SB and MM). Fatty replacement was graded using a five-point scale (0–4)^36^, and STIR signal was classified as positive (+) or negative (-). For each muscle, the MRI slice corresponding to the MSOT acquisition site was identified using anatomical landmarks. For combined analyses, muscles were classified into four imaging categories: T1−/STIR−, T1−/STIR+, T1+/STIR−, T1+/STIR+, representing increasing combinations of structural degeneration and oedema-like changes.

## Statistical analysis

Continuous variables are presented as mean [SD] and categorical variables as counts (percentages). Agreement between repeated acquisitions was assessed using intraclass correlation coefficients (ICC) derived from two-way mixed-effects models with absolute agreement. The same approach was used to evaluate intra-rater and inter-rater reproducibility. ICC values were interpreted according to conventional thresholds^37^. Differences between groups were analysed using two-way ANOVA including group, wavelength (or MSOT parameter) and their interaction, followed by Bonferroni-adjusted post hoc comparisons where appropriate. Similar analyses were performed after stratification according to MRI stage (T1 score, STIR status and combined MRI categories) and Heckmatt grade. Receiver operating characteristic (ROC) analyses assessed the diagnostic performance of MSOT-derived parameters.

To explore abnormalities in radiologically normal muscles, z-scores were calculated for each MSOT parameter relative to muscle-specific healthy control distributions. Values exceeding ± 3 standard deviations were considered outliers.

Associations between MSOT parameters and clinical measures were assessed using Spearman’s correlation coefficients. Patient-level analyses used mean MSOT values averaged across all examined muscles before correlation with CSS, whereas muscle strength was analysed at the individual muscle level. Statistical significance was defined as p < 0.05. Analyses were performed using GraphPad Prism version 9.5.1.

### Standard Protocol Approvals, Registrations, and Patient Consents

The study was registered on ClinicalTrials.gov (NCT05902884). The protocol was approved by the Ethics Committee of Fondazione Policlinico Universitario A. Gemelli IRCCS (Prot. 12392/23; ID 5374) and conducted in accordance with the Declaration of Helsinki. All participants provided written informed consent before enrolment.

## Data Availability

The data that support the findings of this study are available from the corresponding author upon reasonable request and following approval by the institutional Data Protection Officer. Researchers requesting access should provide a brief description of the intended use, including the scientific purpose and the data protection measures to be implemented.

## Results

### Participant characteristics, feasibility and reproducibility

Fifteen patients with genetically confirmed FSHD (13 FSHD1 and 2 FSHD2) and nine healthy volunteers (HVs) were enrolled. Demographic and anthropometric characteristics were comparable between groups (Table 1). Clinical and imaging characteristics of the FSHD cohort are also summarised in Table 1. The examination required a mean of 64 [14] minutes and was well tolerated, with no adverse events reported. MSOT acquisition was successfully completed in all participants, except for one healthy volunteer who stopped the examination early for personal reasons unrelated to the procedure. Overall, 568 muscles were acquired: 360 from patients and 208 from healthy volunteers. Of these, 95% of patient MSOT measurements and 96% of control MSOT measurements had sufficient optoacoustic signal within the ROI and were included in the analysis. Agreement between duplicate acquisitions performed during the same examination was excellent (mean ICC 0.94, 95% Confidence Interval CI 0.938–0.941), with ICC values exceeding 0.80 across all muscles and wavelengths. Accordingly, the average of the two acquisitions was used for subsequent analyses. Repeatability studies performed one week apart demonstrated excellent intra-rater reproducibility (ICC 0.88, 95% CI 0.87–0.89) and similarly excellent inter-rater reproducibility (ICC 0.88, 95% CI 0.87–0.89), confirming the robustness of the acquisition protocol (Figure 1B).

**TABLE 1.** Participant characteristics, muscle imaging findings and muscle strength in FSHD patients. Values are reported as mean ± standard deviation unless otherwise indicated. Demographic and anthropometric variables are shown for healthy volunteers and patients with FSHD, together with *P-*values for between-group differences. FSHD-specific variables include Clinical Severity Scale score and EcoRI fragment length for FSHD type 1. Imaging Characteristics include MRI-based muscle classification according to T1-weighted fatty replacement and STIR hyperintensity. T1+ corresponds to a T1-score > 0. MRI assessment included 300 muscles; biceps brachii and deltoid were outside the MRI acquisition field. RUCT was scored according to Heckmatt grade for muscle echogenicity. RUCT assessment included 350 muscles; 10 muscles were not evaluable because of imaging artifacts. MRI and RUCT categories are reported as number and percentage of assessed muscles. Muscle strength in patients with FSHD was assessed by hand-held dynamometry and is reported in kilograms for each muscle or muscle group. BMI = body mass index; CSS = Clinical Severity Scale; FSHD = facioscapulohumeral muscular dystrophy; HV = healthy volunteers; MRI = magnetic resonance imaging; RUCT = reflected ultrasound computed tomography; STIR = short-tau inversion recovery.

| Participants |  |  |  |
| --- | --- | --- | --- |
|  | HV (N = 9) | FSHD (N = 15) | *P-value* |
| Age (yr) | 47.7 ± 16 | 49.7 ± 13.3 | 0.95 |
| Female sex, n (%) | 5 (56%) | 4 (27%) | 0.21 |
| Height (cm) | 167.4 ± 8.9 | 174 ± 7.8 | 0.06 |
| Weight (kg) | 65.7 ± 16 | 75.8 ± 12.1 | 0.12 |
| BMI (kg/m<sup>2</sup>) | 23.2 ± 4.2 | 24.7 ± 2.6 | 0.22 |
| FSHD |  |  |  |
| FSHD1/FSHD2, n | 13/2 |  |  |
| CSS | 3 ± 2 |  |  |
| EcoRI fragment length (in kb) | 26.9 ± 7 |  |  |
| Imaging Characteristics |  |  |  |
| MRI |  |  |  |
| T1+/STIR+ | n (%) of assessed muscles | 39 (13%) |  |
| T1+/STIR- | n (%) of assessed muscles | 137 (46%) |  |
| T1-/STIR- | n (%) of assessed muscles | 114 (38%) |  |
| T1-/STIR+ | n (%) of assessed muscles | 10 (3%) |  |
| RUCT |  |  |  |
| Heckmatt 1 | n (%) of assessed muscles | 167 (46%) |  |
| Heckmatt 2 | n (%) of assessed muscles | 11 (3%) |  |
| Heckmatt 3 | n (%) of assessed muscles | 17 (5%) |  |
| Heckmatt 4 | n (%) of assessed muscles | 155 (43%) |  |
| Muscle strength (kg) |  |  |  |
| Upper trapezius |  | 5.2 ± 7.6 |  |
| Shoulder abduction |  | 10.5 ± 3.6 |  |
| Elbow flexion |  | 16.8 ± 5.7 |  |
| Knee extension |  | 15.4 ± 8.5 |  |
| Ankle dorsiflexion |  | 6.02 ± 5 |  |
| Knee flexion |  | 12.8 ± 10 |  |
| Ankle plantar flexion |  | 16.5 ± 6.2 |  |

Mean ROI depth did not differ significantly between patients and controls (10.31 [4.51] vs 9.54 [3.83] mm; p = 0.24), indicating that group differences in optoacoustic measurements were unlikely to reflect systematic differences in imaging depth.

### FSHD muscles display a distinct optoacoustic phenotype

When all muscles were analysed together, FSHD muscles differed from healthy muscles at both single wavelength and molecular levels. Single-wavelength signal intensity demonstrated consistently lower optoacoustic profile in FSHD muscles at wavelengths between 700 and 850 nm, whereas signal increased around 930 nm, corresponding to the principal lipid absorption peak within the spectral range of the imaging system (Figure 1C). This spectral profile was accompanied by marked differences in spectrally unmixed parameters. Lipid demonstrated the clearest distinction between groups, with significantly higher values in FSHD muscles than in controls (p < 0.0001). In contrast, collagen-associated signal did not differ significantly between groups. Haemoglobin-related parameters showed evidence of reduced tissue perfusion or oxygenation in FSHD muscles, with lower oxygenated and total haemoglobin compared with healthy controls (Supplementary table 1).

Receiver operating characteristic analysis identified lipid signal as the best-performing MSOT-derived discriminator between FSHD and healthy muscle (Area Under the Curve AUC 0.83 95% CI, 0.662-0.998; p = 0.008), with particularly good performance in clinically relevant muscles including trapezius and medial gastrocnemius^9,38^ (Figure 1D).

Although the overall optoacoustic phenotype was consistent across the cohort, muscle-specific analyses demonstrated greater discrimination in muscles commonly affected in FSHD, whereas relatively spared muscles such as biceps brachii and deltoid showed only limited differences between patients and controls (data not shown).

### MSOT molecular signatures reflect MRI-defined muscle involvement

To investigate the biological significance of the observed optoacoustic phenotype, muscles were stratified according to their appearance on conventional MRI (Figure 2).

**FIGURE 2.**
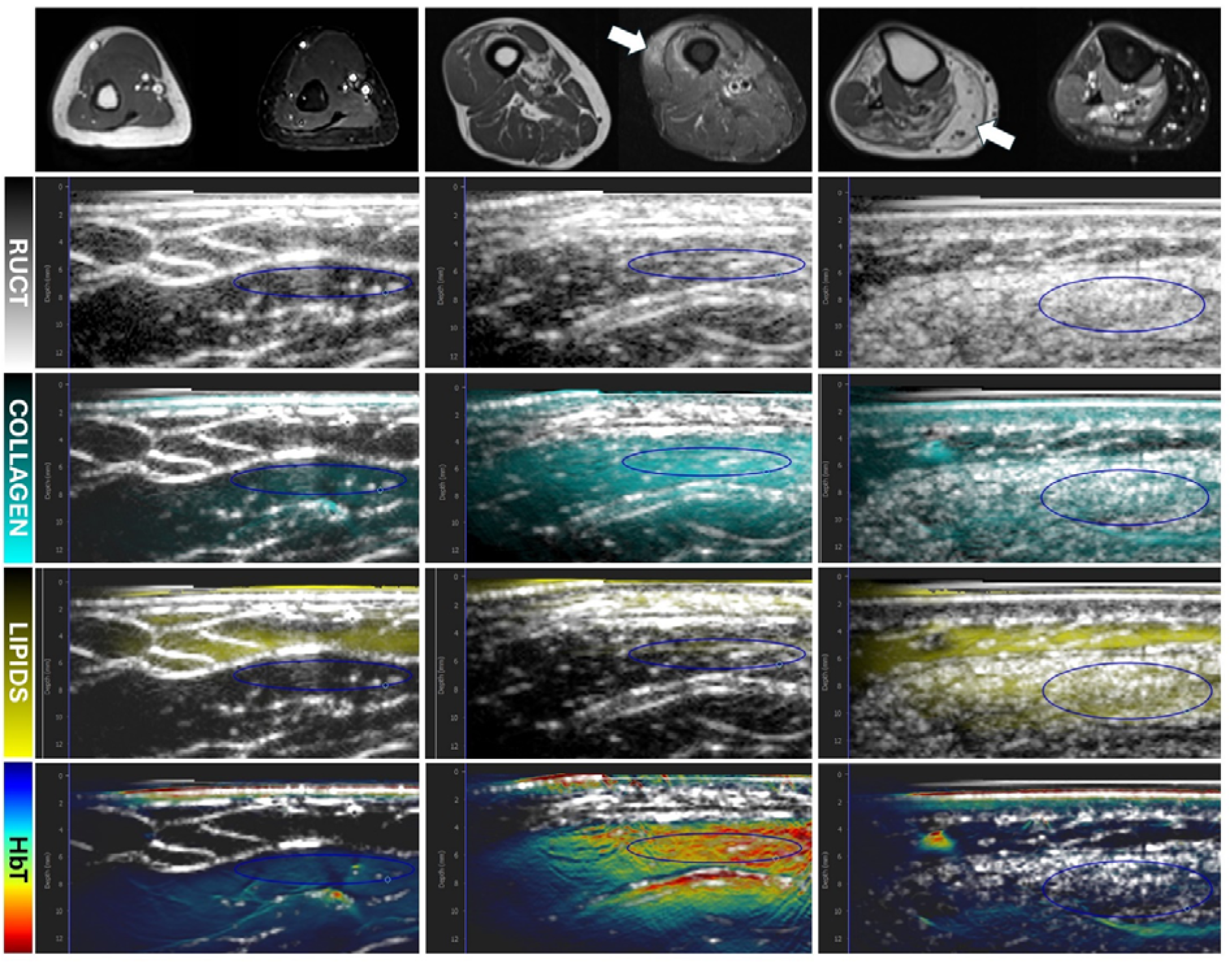
Representative RUCT/MSOT imaging appearances of healthy muscle and different stages of muscle involvement in FSHD. Representative RUCT/MSOT imaging appearances of healthy muscle and different stages of muscle involvement in FSHD. Representative co-registered reflected ultrasound computed tomography (RUCT) and multispectral optoacoustic tomography (MSOT) images illustrate the complementary structural and molecular features of healthy muscle, STIR-positive disease activity and chronic fatty replacement in FSHD. The left column shows the biceps brachii of a healthy control, the middle column shows a distal vastus lateralis muscle from a patient with FSHD classified as T1−/STIR+ on MRI, and the right column shows a medial gastrocnemius muscle from a patient with FSHD classified as T1+/STIR−. MRI images (top row) show T1-weighted and STIR appearances of the corresponding muscles, with arrows indicating the muscles included in the MSOT/RUCT assessment. The biceps brachii image in the control column is an archival MRI image acquired using a separate scanning protocol and is shown for anatomical reference. Blue ellipses indicate the regions of interest (ROIs) used for MSOT quantification. Subsequent rows show RUCT images and MSOT overlays for collagen-associated signal, lipid signal and total haemoglobin (HbT), respectively. The T1−/STIR+ muscle shows increased collagen-associated and HbT signals despite the absence of fatty replacement, whereas the T1+/STIR− muscle shows more prominent lipid signal.

### Fatty replacement

MSOT-derived lipid signal closely reflected MRI-defined fatty infiltration (Figure 3). Lipid signal increased progressively with advancing T1 stage and reached its highest values in severely fatty-replaced muscles (T1 score 3–4 vs. controls and T1 score 0: p < 0.0001; vs. T1 score 1-2: p = 0.0062; Supplementary table 2). By contrast, collagen-associated signal remained relatively stable across T1 categories. Haemoglobin-related parameters demonstrated the opposite trend. Oxygenated haemoglobin, total haemoglobin and tissue oxygen saturation progressively declined with increasing fatty replacement, suggesting that advanced structural degeneration is accompanied by loss of metabolically active and vascularised muscle tissue. Collectively, these findings demonstrate that the principal molecular signature of chronic muscle degeneration in FSHD consists of increasing lipid content together with progressive reduction of haemoglobin-related signals.

**FIGURE 3.**
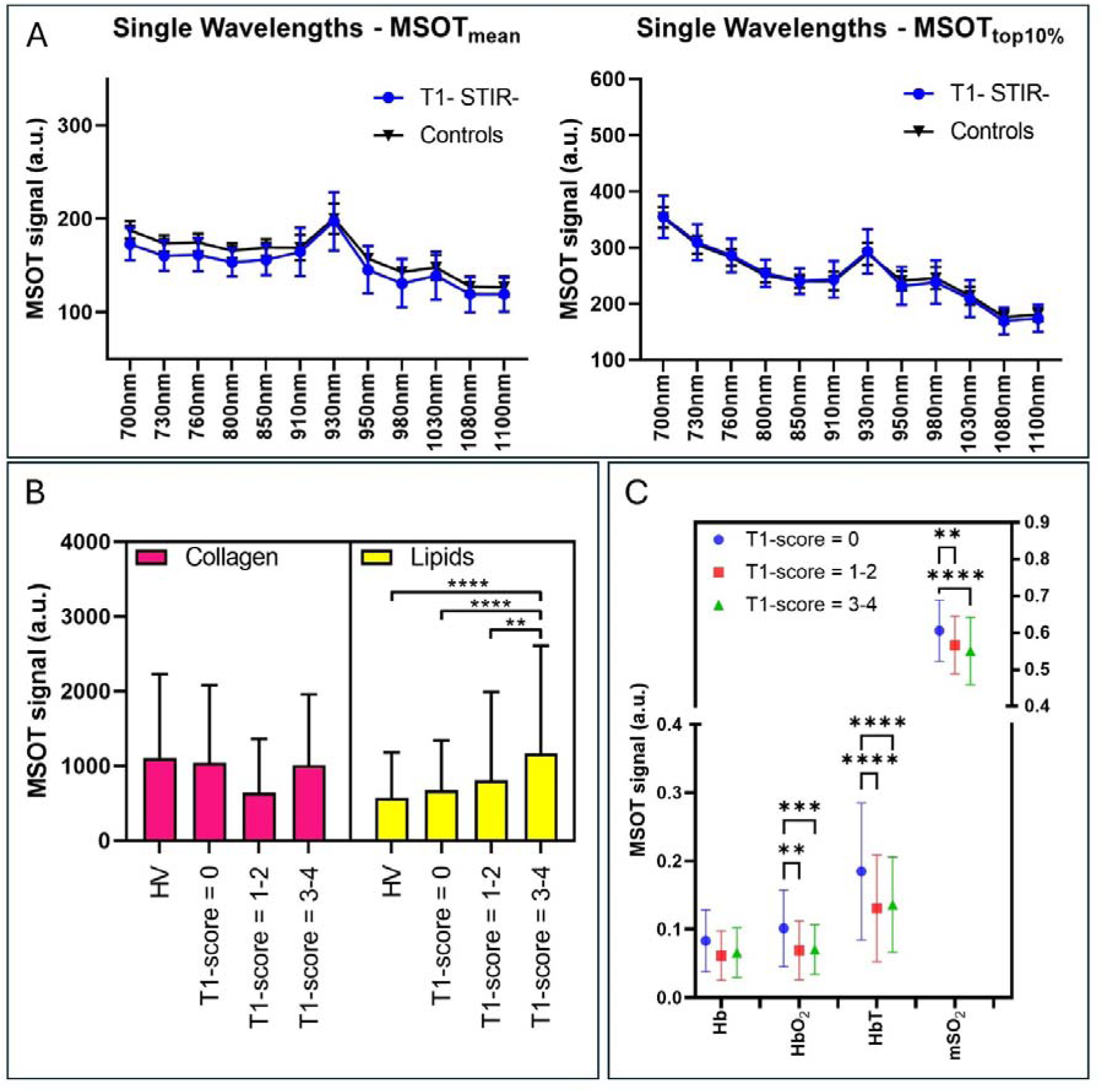
MSOT molecular signatures reflect progressive fatty replacement. (**A**) MSOT_mean_ and MSOT_top10%_ single-wavelength signal intensity in T1-/STIR-patient muscles and control muscles demonstrate broadly preserved optoacoustic profiles in structurally normal muscles. (**B**) Collagen-associated and lipid signals stratified by T1 score. Lipid signal increased with fat replacement and was highest in severely affected muscles, whereas collagen-associated signal remained relatively stable across T1 categories. (**C**) Haemoglobin-related parameters and tissue oxygen saturation across T1 categories. Increasing fat replacement was associated with lower oxygenated haemoglobin (HbO□), total haemoglobin (HbT) and tissue oxygen saturation (mSO□). ** p < 0.01, *** p < 0.001, **** p < 0.0001.

### STIR-positive muscles exhibit a distinct molecular phenotype

Unlike chronic fatty replacement, STIR positivity was associated with a qualitatively different optoacoustic profile (Figure 4). Compared with STIR-negative muscles, STIR-positive muscles demonstrated an upward shift of the optoacoustic spectrum, particularly at longer wavelengths. Spectral unmixing revealed significantly higher collagen-associated (MSOT_top10%_ collagen 2594 [1801] vs 1709 [1553] a.u.; p = 0.0012) and lipid signals (3332 [2512] vs 2351 [2088] a.u.; p = 0.0004) together with increased deoxygenated haemoglobin and total haemoglobin (Supplementary table 3). Among the subgroup of muscles that were STIR-positive but showed no fatty replacement (T1−/STIR+), collagen-associated and haemoglobin signals were already elevated despite preserved structural appearance on T1-weighted MRI (Figure 2). These observations indicate that STIR-positive muscles are characterised by a distinct molecular signature suggestive of active tissue remodelling.

**FIGURE 4.**
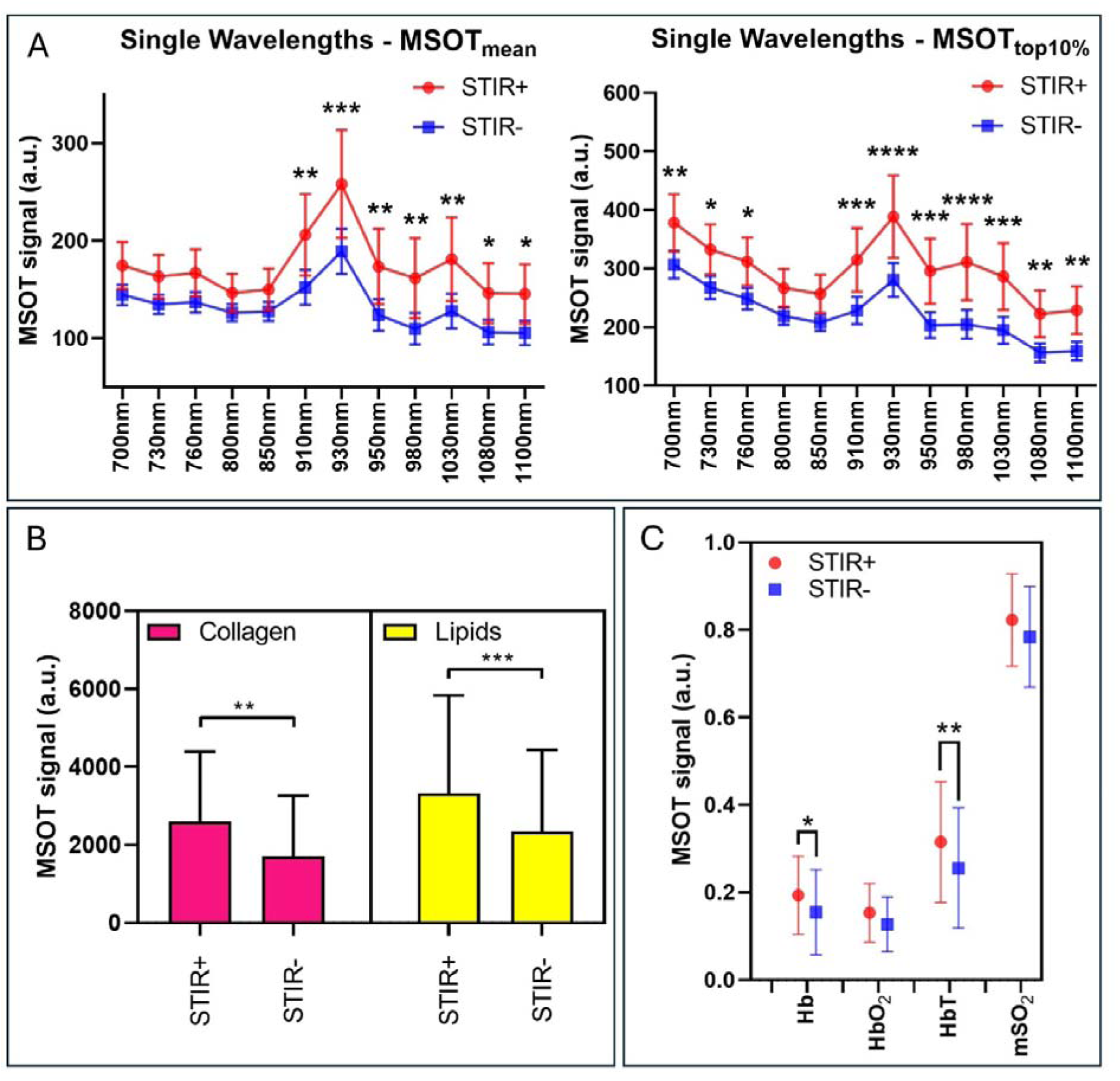
STIR-positive muscles display a distinct optoacoustic molecular phenotype. (**A**) MSOT_mean_ and MSOT_top10%_ single-wavelength signal intensity in STIR+ and STIR-muscles. STIR+ muscles showed an upward shift of the optoacoustic spectrum. (**B**) MSOT_top10%_ collagen-associated and lipid signals stratified by STIR status. STIR-positive muscles exhibited higher collagen-associated and lipid signals than STIR-negative muscles. (**C**) MSOT_top10%_ haemoglobin-related parameters and tissue oxygen saturation according to STIR status. STIR-positive muscles demonstrated increased deoxygenated haemoglobin (Hb) and total haemoglobin (HbT). Hb = deoxygenated haemoglobin; HbO2 = oxygenated haemoglobin; HbT = total haemoglobin; mSO2 = tissue oxygen saturation. * p < 0.05, ** p < 0.01, *** p < 0.001, **** p < 0.0001.

### Relationship between MSOT and RUCT

MSOT findings only partially overlapped with RUCT-derived Heckmatt grading (Figure 5). Muscles classified as Heckmatt grade 1 showed optoacoustic spectra comparable to those of healthy controls. In contrast, grade 4 muscles demonstrated increased signal around lipid-related wavelengths together with markedly increased lipid signal. Intermediate Heckmatt grades exhibited a less linear optoacoustic profile characterised by reduced signal intensity across much of the spectrum. Unlike lipid signal, collagen-associated signal showed no clear relationship with Heckmatt grade. Haemoglobin-related parameters were reduced in muscles with increasing ultrasound abnormalities (Supplementary table 4). Overall, these findings suggest that RUCT and MSOT interrogate different aspects of muscle pathology and therefore provide complementary rather than redundant information.

**FIGURE 5.**
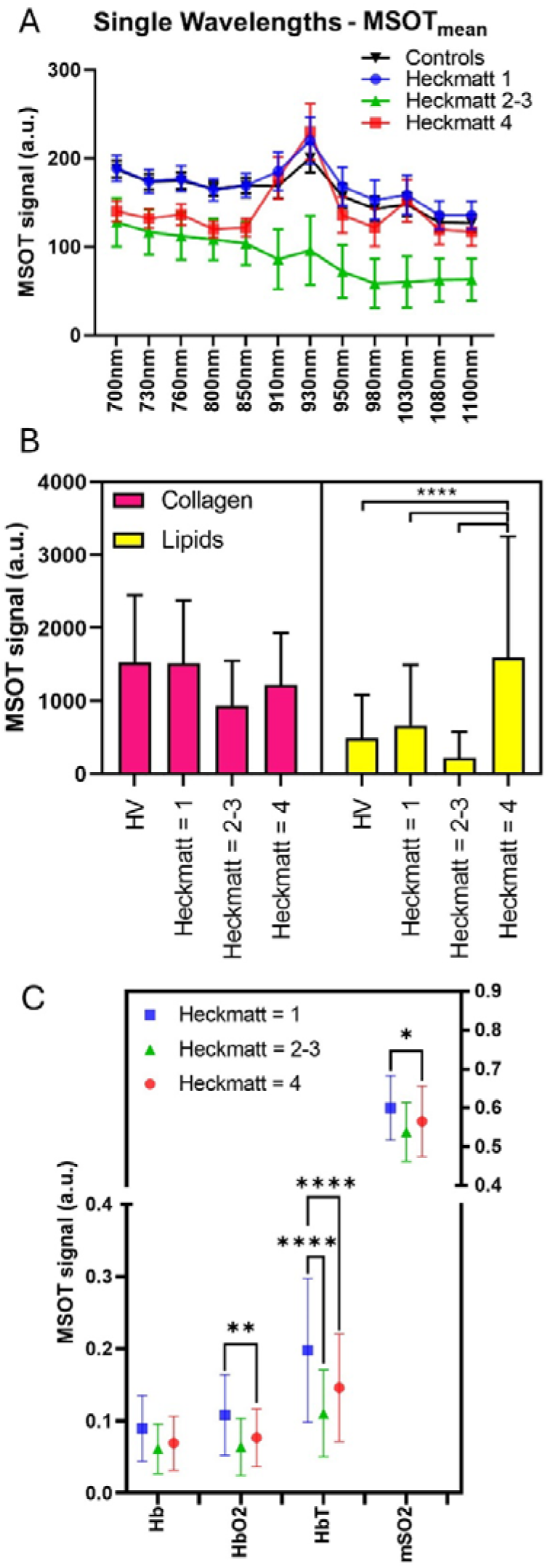
MSOT provides complementary information to ultrasound-derived muscle architecture. (**A**) MSOT_mean_ single-wavelength signal intensity in control muscles and patient muscles stratified by Heckmatt grade. Muscles with intermediate ultrasound abnormalities (Heckmatt grades 2-3) displayed a non-linear reduction in optoacoustic intensity, whereas severely affected muscles (grade 4) showed higher signal around lipid-related wavelengths. (**B**) Collagen-associated and lipid signals by Heckmatt category. Lipid signal increased in grade 4 muscles, whereas collagen-associated signal showed no clear relationship with ultrasound grade. (**C**) Haemoglobin-related parameters and tissue oxygen saturation by Heckmatt category. Increasing ultrasound abnormalities were associated with lower haemoglobin-related parameters, consistent with progressive loss of metabolically active muscle tissue. Hb = deoxygenated haemoglobin; HbO2 = oxygenated haemoglobin; HbT = total haemoglobin; mSO2 = tissue oxygen saturation. * p < 0.05, ** p < 0.01, **** p < 0.0001.

### Isolated MSOT abnormalities are detectable in normal muscles on conventional imaging

At the group level, muscles classified as normal by both MRI and RUCT exhibited optoacoustic spectra that were largely indistinguishable from those of healthy controls. However, analysis at the individual muscle level revealed a different pattern. Thirteen of 141 radiologically normal muscles (9%), originating from six patients, demonstrated at least one MSOT-derived parameter outside the predefined reference range (Figure 6). The most frequent abnormality was increased lipid signal, followed by altered tissue oxygen saturation. This data demonstrates a degree of biological heterogeneity among conventionally normal muscles and suggests that MSOT may detect molecular alterations that remain below the detection threshold of structural imaging.

**FIGURE 6.**
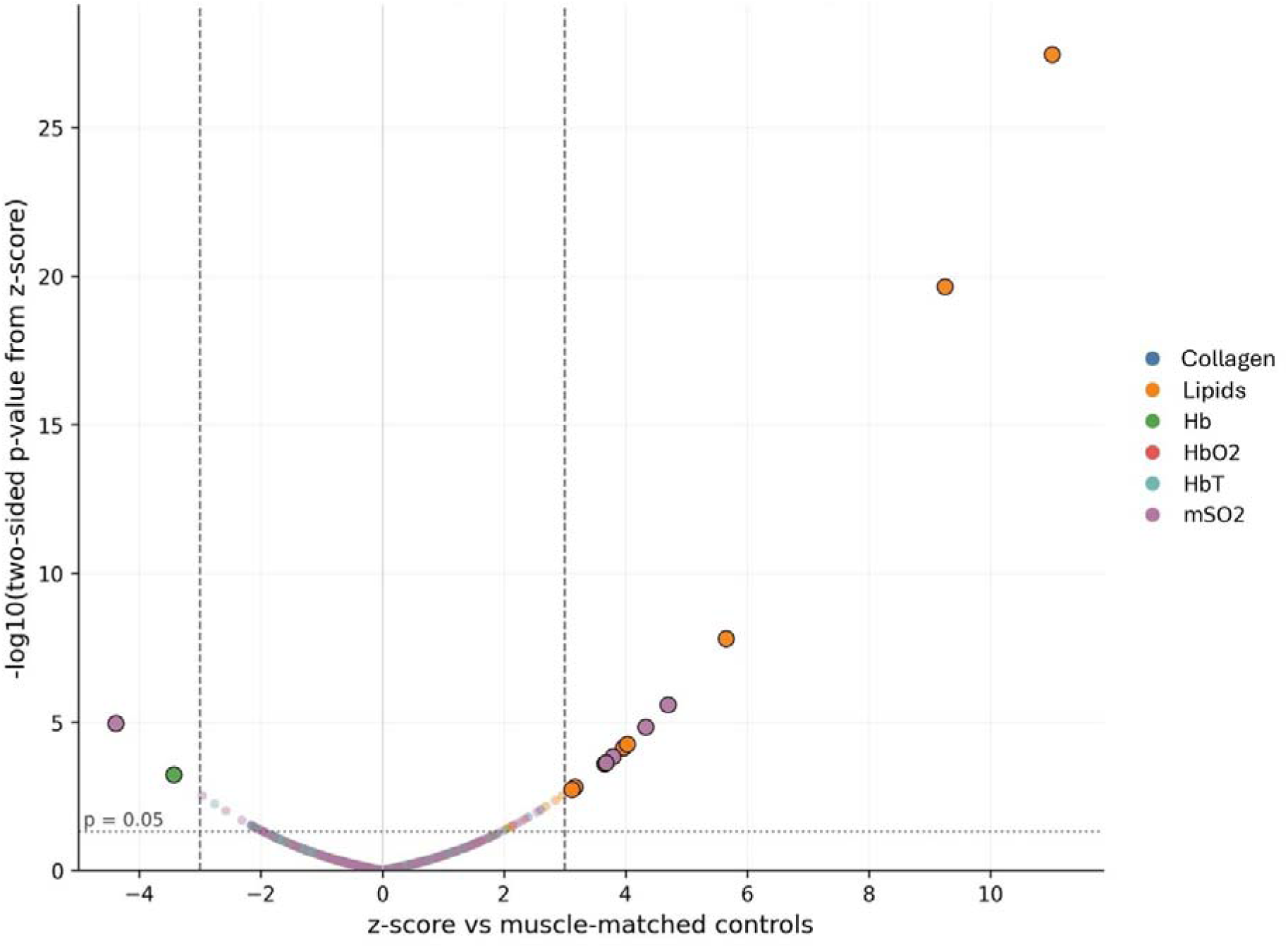
Individual muscle analysis identifies isolated MSOT abnormalities in radiologically normal muscles. Each point represents one optoacoustic parameter in one patient-muscle classified as radiologically normal. The x-axis shows the z-score relative to the muscle-matched healthy control distribution; the y-axis shows-log10 of the two-sided *P* value derived from the z-score. Vertical dashed lines indicate z-score thresholds of-3 and +3, and the horizontal dotted line indicates *P* = 0.05. Most radiologically normal muscles clustered within the control distribution, whereas selected muscles showed outlying optoacoustic values, most frequently increased lipid signal (seven muscles from three patients). Hb = deoxygenated haemoglobin; HbO2 = oxygenated haemoglobin; HbT = total haemoglobin; mSO2 = tissue oxygen saturation.

### Clinical associations

MSOT-derived parameters demonstrated modest but biologically consistent associations with clinical measures. At the individual muscle level, increased lipid signal correlated with lower muscle strength (r =-0.27; p < 0.001), whereas haemoglobin-related parameters correlated positively with force generation (deoxygenated haemoglobin r = 0.20, p = 0.004; total haemoglobin r = 0.20, p = 0.004). Similar associations were observed for shorter-wavelength spectral signals. At the patient level, lower average oxygenated haemoglobin and total haemoglobin signals correlated with greater disease severity as measured by the CSS (r =-0.60, p = 0.02 and r =-0.60, p = 0.02 respectively). No significant associations were observed between disease severity and collagen-associated or lipid signals averaged across muscles (Supplementary table 5). Taken together, these findings indicate that MSOT-derived molecular measures may reflect local muscle dysfunction and, to some extent, overall disease burden in FSHD.

## Discussion

In this pilot study, we demonstrate that multispectral optoacoustic tomography is feasible, reproducible and well tolerated in patients with FSHD, while providing molecular information that complements conventional muscle imaging. Rather than simply reproducing structural abnormalities detected by MRI, MSOT identified distinct optoacoustic signatures associated with different stages of muscle involvement, including chronic fatty replacement and disease activity, and showed correlation with functional impairment. Importantly, a subset of muscles that appeared normal on conventional imaging displayed isolated MSOT abnormalities, suggesting that advanced optoacoustic imaging may reveal biological heterogeneity not captured by current conventional techniques.

The excellent reproducibility observed across repeated acquisitions and between operators represents an important prerequisite for any imaging biomarker intended for longitudinal studies or clinical trials. FSHD poses particular challenges because of its marked spatial heterogeneity, variable disease progression and selective involvement of individual muscles^4,8,9,39^. Despite these challenges, MSOT measurements remained highly reproducible across multiple anatomical regions, supporting the technical robustness of the acquisition protocol^40^. Although the examination currently requires approximately one hour, further optimisation of acquisition workflows and automated image analysis could improve its practicality for future, multicentre studies.

The close relationship between MSOT-derived lipid signal and MRI-defined fatty replacement provides important orthogonal validation of the technique. The characteristic reduction in shorter-wavelength optoacoustic signals together with increased signal around 930 nm was accompanied by a marked increase in spectrally unmixed lipid signal, consistent with the optical absorption properties of lipid within the wavelength range of the imaging system^41^. Importantly, lipid signal continued to distinguish severely affected muscles, at variance with increased echogenicity on ultrasound^22,42^, suggesting that MSOT remains informative even in advanced disease. Nevertheless, we do not consider this the principal strength of the technique. MRI already provides an excellent assessment of fatty replacement, particularly when quantitative Dixon imaging is available. Rather, the concordance between MRI and MSOT demonstrates that optoacoustic imaging accurately captures one established component of muscle pathology while simultaneously providing additional accurate molecular information.

The findings observed in STIR-positive muscles are likely the most biologically informative aspect of this study. Whereas chronic fatty replacement was primarily characterised by increasing lipid signal and declining haemoglobin-related parameters, in keeping with capillary bed depletion documented in FSHD muscle biopsies^19,43^, STIR-positive muscles exhibited a distinct molecular profile combining increased collagen-associated signal, elevated lipid signal and increased deoxygenated and total haemoglobin. This pattern is compatible with active tissue remodelling, during which extracellular matrix turnover, early lipid accumulation and vascular alterations coexist before complete structural degeneration has occurred. Although the precise biological substrates of these signals require further investigation, the observations are consistent with previous MRI-guided biopsy studies demonstrating inflammatory infiltrates, increased fibrosis and vessel proliferation, possibly directly linked to DUX4 target gene activation, within STIR-positive FSHD muscles^14–16,18,44^.

In particular, the haemoglobin-related findings provide an additional dimension to the biological information obtained by MSOT. Reduced oxygenated and total haemoglobin in chronically affected muscles, together with their association with greater disease severity, are consistent with previous observations suggesting altered muscle perfusion and vascular physiology in FSHD^20^. Conversely, the relative increase in deoxygenated and total haemoglobin observed in STIR-positive muscles may reflect increased vascular volume and altered oxygen extraction accompanying active disease. Together, these findings suggest that MSOT interrogates physiological processes that extend beyond structural tissue composition and may provide a more comprehensive characterisation of biologically active muscle than conventional imaging alone.

Particular caution is warranted when interpreting the collagen-associated signal. The spectral signatures of collagen and lipid partially overlap, and current spectral unmixing algorithms cannot completely separate all endogenous absorbers under every biological condition^45^. Consequently, the collagen-associated signal reported here should not be interpreted as a direct measure of histological fibrosis. Rather, it represents an optoacoustic biomarker influenced by collagen-rich extracellular matrix that requires further validation against quantitative histopathology. Recent advances in image reconstruction, spectral unmixing and model-based algorithms are likely to improve the specificity and robustness of these measurements in future studies^46,47^. Nevertheless, the observation that collagen-associated optoacoustic readout was preferentially increased in STIR-positive rather than chronically fatty muscles is intriguing and consistent with the concept that extracellular matrix remodelling is a dynamic process accompanying disease activity rather than simply reflecting end-stage fibrosis^16^.

One of the more interesting findings was the identification of isolated optoacoustic abnormalities in approximately 9% of muscles that appeared normal on both MRI and RUCT. At the group level these muscles were indistinguishable from healthy controls. However, the presence of individual outliers suggests biological heterogeneity among structurally “normal” muscles. These observations support the testable hypothesis that molecular alterations may precede, accompany or evolve independently from the structural changes detectable by current imaging techniques. Similar observations have been reported in Pompe disease, where optoacoustic abnormalities extended beyond conventional imaging findings^28^. Whether these isolated abnormalities predict disease evolution in terms of subsequent STIR positivity, fatty replacement or functional decline can only be determined through longitudinal follow-up.

Taken together, our findings support a conceptual distinction between structural and molecular imaging biomarkers in FSHD. Conventional MRI provides an excellent framework for staging structural muscle degeneration through assessment of fatty replacement and oedema-like changes. Similarly, RUCT reflects alterations in muscle architecture and echogenicity^48^. MSOT offers complementary molecular information related to tissue composition, extracellular matrix remodelling and haemoglobin dynamics. We therefore envisage MSOT as an adjunctive imaging modality capable of enriching the biological interpretation of conventional imaging rather than competing with it. This complementary role may become particularly valuable in therapeutic trials, where biomarkers reflecting active tissue remodelling could provide information distinct from the relatively slow structural changes observed on conventional imaging.

Several limitations should be acknowledged. This was a single-centre pilot study with a relatively small sample size, limiting statistical power for subgroup analyses. MRI assessment relied on semiquantitative visual grading rather than quantitative Dixon fat fraction or T2 mapping, although this approach reflects routine clinical practice and allowed direct comparison with established imaging staging systems^8^. The current implementation of MSOT is restricted to relatively superficial muscles and does not compensate for depth-dependent optical attenuation, factors that may influence quantitative measurements. Finally, the cross-sectional design precludes conclusions regarding temporal changes or prognostic value.

These limitations also define the next steps for the field. Longitudinal studies integrating MSOT with quantitative MRI, circulating biomarkers and MRI-guided muscle biopsies will be required to determine the biological specificity, responsiveness and prognostic value of optoacoustic measures. Future studies should also investigate whether combining MSOT with standardized physiological challenges, such as exercise, provides additional functional information by revealing dynamic haemoglobin and tissue oxygenation responses beyond resting measurements^49^. Such longitudinal and multimodal approaches will help determine whether MSOT-derived molecular signatures identify muscles undergoing active remodelling, predict subsequent structural progression or serve as pharmacodynamic biomarkers in therapeutic trials.

In conclusion, this study demonstrates that multispectral optoacoustic tomography is a feasible and reproducible imaging modality that provides complementary molecular information on muscle involvement in FSHD. Distinct optoacoustic signatures were associated with chronic fatty replacement and STIR-positive areas of disease activity, while isolated abnormalities in normal muscles on conventional radiology suggest sensitivity to biological heterogeneity beyond structural imaging. These findings support further development of MSOT as a multimodal imaging biomarker and provide the foundation for future longitudinal studies evaluating its role in monitoring disease activity and therapeutic response in FSHD.

## Acknowledgements

We are extremely grateful to the FSHD Italia Onlus association, the patients and their families for their constant support. A large language model (ChatGPT, GPT 5.5, OpenAI) was used to improve the clarity and conciseness of specific sentences in the manuscript, and to generate schematic diagrams in Figure 1A. All suggestions were critically reviewed and revised by the authors to ensure the accuracy and integrity of the scientific content.

## References

1. Deenen JCW, Arnts H, van der Maarel SM, et al. Population-based incidence and prevalence of facioscapulohumeral dystrophy. Neurology. 2014;83(12):1056–1059. doi:10.1212/WNL.0000000000000797

2. Lemmers RJLF, van der Vliet PJ, Klooster R, et al. A unifying genetic model for facioscapulohumeral muscular dystrophy. Science. 2010;329(5999):1650–1653. doi:10.1126/science.1189044

3. Tihaya MS, Mul K, Balog J, et al. Facioscapulohumeral muscular dystrophy: the road to targeted therapies. Nat Rev Neurol. 2023;19(2):91–108. doi:10.1038/s41582-022-00762-2

4. Ruggiero L, Mele F, Manganelli F, et al. Phenotypic Variability Among Patients With D4Z4 Reduced Allele Facioscapulohumeral Muscular Dystrophy. JAMA Netw Open. 2020;3(5):e204040. doi:10.1001/jamanetworkopen.2020.4040

5. Ricci G, Scionti I, Sera F, et al. Large scale genotype–phenotype analyses indicate that novel prognostic tools are required for families with facioscapulohumeral muscular dystrophy. Brain. 2013;136(11):3408–3417. doi:10.1093/brain/awt226

6. Torchia E, Vandeputte P, Monforte M, et al. Camptocormia as a Phenotypic Variant of FSHD in the Elderly: Clinical, Genetic, and Imaging Features. Eur J Neurol. 2025;32(10):e70332. doi:10.1111/ene.70332

7. Tammam G, Dhifallah S, Yang H, et al. Late-onset facioscapulohumeral muscular dystrophy defines a distinct clinical subgroup. Neuromuscul Disord. 2026;62:106410. doi:10.1016/j.nmd.2026.106410

8. Monforte M, Laschena F, Ottaviani P, et al. Tracking muscle wasting and disease activity in facioscapulohumeral muscular dystrophy by qualitative longitudinal imaging. J Cachexia Sarcopenia Muscle. Published online October 30, 2019:jcsm.12473. doi:10.1002/jcsm.12473

9. Tasca G, Monforte M, Ottaviani P, et al. Magnetic Resonance Imaging in a large cohort of facioscapulohumeral muscular dystrophy patients: pattern refinement and implications for clinical trials. Ann Neurol. Published online March 19, 2016. doi:10.1002/ana.24640

10. Tasca G, Monforte M, Iannaccone E, et al. Upper girdle imaging in facioscapulohumeral muscular dystrophy. PloS One. 2014;9(6):e100292. doi:10.1371/journal.pone.0100292

11. Gerevini S, Scarlato M, Maggi L, et al. Muscle MRI findings in facioscapulohumeral muscular dystrophy. Eur Radiol. 2016;26(3):693–705. doi:10.1007/s00330-015-3890-1

12. Vincenten SC, Teeselink S, Mul K, et al. Muscle imaging in facioscapulohumeral muscular dystrophy research: A scoping review and expert recommendations. Neuromuscul Disord. 2025;47:105274. doi:10.1016/j.nmd.2025.105274

13. Monforte M, Attarian S, Vissing J, Diaz-Manera J, Tasca G. 265th ENMC international workshop: Muscle imaging in Facioscapulohumeral Muscular Dystrophy (FSHD): Relevance for clinical trials. 22–24 April 2022, Hoofddorp, The Netherlands. Neuromuscul Disord. Published online October 2022:S096089662200699X. doi:10.1016/j.nmd.2022.10.005

14. Di Pietro L, Giacalone F, Ragozzino E, et al. Non-myogenic mesenchymal cells contribute to muscle degeneration in facioscapulohumeral muscular dystrophy patients. Cell Death Dis. 2022;13(9):793. doi:10.1038/s41419-022-05233-6

15. Jung U, Wei E, Ahsan H, et al. Matrix metalloproteinases are hallmark early biomarkers and therapeutic targets in FSHD. JCI Insight. 10(21):e195104. doi:10.1172/jci.insight.195104

16. Ragozzino E, Bortolani S, Di Pietro L, et al. Muscle fibrosis as a prognostic biomarker in facioscapulohumeral muscular dystrophy: a retrospective cohort study. Acta Neuropathol Commun. 2023;11(1):165. doi:10.1186/s40478-023-01660-4

17. Tasca G, Pescatori M, Monforte M, et al. Different molecular signatures in magnetic resonance imaging-staged facioscapulohumeral muscular dystrophy muscles. PloS One. 2012;7(6):e38779. doi:10.1371/journal.pone.0038779

18. Osborne RJ, Welle S, Venance SL, Thornton CA, Tawil R. Expression profile of FSHD supports a link between retinal vasculopathy and muscular dystrophy. Neurology. 2007;68(8):569–577. doi:10.1212/01.wnl.0000251269.31442.d9

19. Statland JM, Odrzywolski KJ, Shah B, et al. Immunohistochemical Characterization of FacioscapulohumeralMuscular Dystrophy Muscle Biopsies. J Neuromuscul Dis. 2015;2(3):291–299. doi:10.3233/JND-150077

20. Olivier N, Boissière J, Allart E, et al. Evaluation of muscle oxygenation by near infrared spectroscopy in patients with facioscapulohumeral muscular dystrophy. Neuromuscul Disord. 2016;26(1):47–55. doi:10.1016/j.nmd.2015.10.004

21. Mul K, Horlings CGC, Vincenten SCC, Voermans NC, van Engelen BGM, van Alfen N. Quantitative muscle MRI and ultrasound for facioscapulohumeral muscular dystrophy: complementary imaging biomarkers. J Neurol. 2018;265(11):2646–2655. doi:10.1007/s00415-018-9037-y

22. Vincenten SCC, Teeselink S, Voermans NC, van Engelen BGM, Mul K, van Alfen N. Establishing the role of muscle ultrasound as an imaging biomarker in facioscapulohumeral muscular dystrophy. Neuromuscul Disord. 2023;33(12):936–944. doi:10.1016/j.nmd.2023.10.015

23. Wijntjes J, Saris C, Doorduin J, van Alfen N, van Engelen B, Mul K. Improving Heckmatt muscle ultrasound grading scale through Rasch analysis. Neuromuscul Disord. 2024;42:14–21. doi:10.1016/j.nmd.2024.07.001

24. Knieling F, Lee S, Ntziachristos V. A primer on current status and future opportunities of clinical optoacoustic imaging. Npj Imaging. 2025;3(1):4. doi:10.1038/s44303-024-00065-9

25. Tan L, Meyer S, Zschüntzsch J, Rother U, Knieling F. Optoacoustic muscle imaging. J Neuromuscul Dis. Published online April 27, 2026:22143602261443594. doi:10.1177/22143602261443594

26. Regensburger AP, Fonteyne LM, Jüngert J, et al. Detection of collagens by multispectral optoacoustic tomography as an imaging biomarker for Duchenne muscular dystrophy. Nat Med. Published online December 2, 2019. doi:10.1038/s41591-019-0669-y

27. Regensburger AP, Wagner AL, Danko V, et al. Multispectral optoacoustic tomography for non-invasive disease phenotyping in pediatric spinal muscular atrophy patients. Photoacoustics. 2021;25:100315. doi:10.1016/j.pacs.2021.100315

28. Tan L, Zschüntzsch J, Meyer S, et al. Non-invasive optoacoustic imaging of glycogen-storage and muscle degeneration in late-onset Pompe disease. Nat Commun. 2024;15(1):7843. doi:10.1038/s41467-024-52143-6

29. Günther JS, Knieling F, Träger AP, et al. Targeting Muscular Hemoglobin Content for Classification of Peripheral Arterial Disease by Noninvasive Multispectral Optoacoustic Tomography. JACC Cardiovasc Imaging. 2023;16(5):719–721. doi:10.1016/j.jcmg.2022.11.010

30. Wittig T, Winther B, Reichl C, Schmidt A, Scheinert D, Steiner S. Optoacoustic imaging in lower extremity revascularization: A novel technique to assess perioperative muscle perfusion. Photoacoustics. 2025;45:100756. doi:10.1016/j.pacs.2025.100756

31. Knieling F, Neufert C, Hartmann A, et al. Multispectral Optoacoustic Tomography for Assessment of Crohn’s Disease Activity. N Engl J Med. 2017;376(13):1292–1294. doi:10.1056/NEJMc1612455

32. Hallasch S, Giese N, Stoffels I, Klode J, Sondermann W. Multispectral optoacoustic tomography might be a helpful tool for noninvasive early diagnosis of psoriatic arthritis. Photoacoustics. 2020;21:100225. doi:10.1016/j.pacs.2020.100225

33. Tascilar K, Fagni F, Kleyer A, et al. Non-invasive metabolic profiling of inflammation in joints and entheses by multispectral optoacoustic tomography. Rheumatology. 2023;62(2):841–849. doi:10.1093/rheumatology/keac346

34. Ricci E, Galluzzi G, Deidda G, et al. Progress in the molecular diagnosis of facioscapulohumeral muscular dystrophy and correlation between the number of KpnI repeats at the 4q35 locus and clinical phenotype. Ann Neurol. 1999;45(6):751–757.

35. Tasca G, Monforte M, Iannaccone E, et al. Muscle MRI in female carriers of dystrophinopathy. Eur J Neurol. 2012;19(9):1256–1260. doi:10.1111/j.1468-1331.2012.03753.x

36. Fischer D, Kley RA, Strach K, et al. Distinct muscle imaging patterns in myofibrillar myopathies. Neurology. 2008;71(10):758–765. doi:10.1212/01.wnl.0000324927.28817.9b

37. Landis JR, Koch GG. The Measurement of Observer Agreement for Categorical Data. Biometrics. 1977;33(1):159–174. doi:10.2307/2529310

38. Monforte M, Bortolani S, Torchia E, et al. Diagnostic magnetic resonance imaging biomarkers for facioscapulohumeral muscular dystrophy identified by machine learning. J Neurol. 2022;269(4):2055–2063. doi:10.1007/s00415-021-10786-1

39. Katz NK, Hogan J, Delbango R, Cernik C, Tawil R, Statland JM. Predictors of functional outcomes in patients with facioscapulohumeral muscular dystrophy. Brain. 2021;144(11):3451–3460. doi:10.1093/brain/awab326

40. Wagner AL, Danko V, Federle A, et al. Precision of handheld multispectral optoacoustic tomography for muscle imaging. Photoacoustics. 2021;21:100220. doi:10.1016/j.pacs.2020.100220

41. Graham MT, Sharma A, Padovano WM, et al. Optical absorption spectra and corresponding in vivo photoacoustic visualization of exposed peripheral nerves. J Biomed Opt. 2023;28(9):097001. doi:10.1117/1.JBO.28.9.097001

42. Wijntjes J, van der Hoeven J, Saris CGJ, Doorduin J, van Alfen N. Visual versus quantitative analysis of muscle ultrasound in neuromuscular disease. Muscle Nerve. 2022;66(3):253–261. doi:10.1002/mus.27669

43. Greco A, Kusters B, Mann R, et al. Immunohistological and electron microscopy profile of unique TIRM-MRI guided muscle biopsies of FSHD patients. J Neuromuscul Dis. Published online October 6, 2025:22143602251380426. doi:10.1177/22143602251380426

44. Frisullo G, Frusciante R, Nociti V, et al. CD8(+) T cells in facioscapulohumeral muscular dystrophy patients with inflammatory features at muscle MRI. J Clin Immunol. 2011;31(2):155–166. doi:10.1007/s10875-010-9474-6

45. Park E, Lee YJ, Lee C, Eom TJ. Effective photoacoustic absorption spectrum for collagen-based tissue imaging. J Biomed Opt. 2020;25(5):056002. doi:10.1117/1.JBO.25.5.056002

46. Di Giacinto F, Riente A, Mignini I, et al. Advancing multispectral optoacoustic tomography (MSOT): Phasor analysis for real-time spectral unmixing. Comput Biol Med. 2025;195:110586. doi:10.1016/j.compbiomed.2025.110586

47. Assi H, Cao R, Castelino M, et al. A review of a strategic roadmapping exercise to advance clinical translation of photoacoustic imaging: From current barriers to future adoption. Photoacoustics. 2023;32:100539. doi:10.1016/j.pacs.2023.100539

48. Danko V, Jüngert J, Schuessler S, et al. Hybrid reflected-ultrasound computed tomography versus B-mode-ultrasound for muscle scoring in spinal muscular atrophy. J Neuroimaging. 2023;33(3):393–403. doi:10.1111/jon.13081

49. Caranovic M, Kempf J, Li Y, et al. Derivation and validation of a non-invasive optoacoustic imaging biomarker for detection of patients with intermittent claudication. Commun Med. 2025;5:88. doi:10.1038/s43856-025-00801-1

