## Supplementary document for "Optoacoustic molecular signatures of muscle involvement in facioscapulohumeral dystrophy compared to conventional imaging"

| Patient position | Anatomical muscle site | Landmark description |
|---|---|---|
| Supine position | *Deltoid* | 1/2 between the lateral edge of the acromion and the deltoid tuberosity |
| | *Biceps brachii* | 2/3 between the acromion and the antecubital fossa |
| | *Recti abdominis*<br>- Upper part | At the level of $10^{th}$ rib |
| | - Lower part | 1/3 between the anterior superior iliac spine and the umbilicus |
| | *Rectus femoris* | 1/2 between anterior superior iliac spine and superior tip of patella |
| | *Vastus lateralis*<br>- Proximal | 1/2 between anterior superior iliac spine and superior border of the patella, at the same level as the rectus femoris measurement site, displaced laterally. |
| | - Distal | 3/4 between anterior superior iliac spine and superior tip of patella |
| | *Tibialis anterior* | 1/4 between the fibular head and the lateral malleolus |
| Prone position | *Paraspinal* | Immediately below the costal margin, adjacent to lumbar spinous processes. |
| | *Biceps femoris* | 1/2 between lateral femoral condyle and the greater trochanter |
| | *Medial gastrocnemius* | 1/3 between the fibular head and medial malleolus |
| Sitting position | *Trapezius* | ½ between the acromion and the spinous process of C7 vertebra (at the level of C7, aligned with the mid-clavicular line) |

**Standardized anatomical landmarks for MSOT acquisition.**
Probe placement sites used for MSOT acquisition are reported for each muscle. Anatomical landmarks were used to standardize bilateral assessment of upper-limb, trunk and lower-limb muscles across participants.
