## Supplementary Table 1 for "Optoacoustic molecular signatures of muscle involvement in facioscapulohumeral dystrophy compared to conventional imaging"

| | | FSHD | | Controls | | | FSHD | Controls | FSHD | Controls |
|---|---|---|---|---|---|---|---|---|---|---|
| | | *Mean* | *SD* | *Mean* | *SD* | *P-value* | *N* | *N* | % (out of 360) | % (out of 208) |
| SWL mean | 700nm | 162,03 | 87,52 | 188,00 | 100,41 | **0,0003** | 350 | 202 | 97 | 97 |
| | 730nm | 150,46 | 80,35 | 173,54 | 92,60 | **0,0014** | 350 | 202 | 97 | 97 |
| | 760nm | 153,45 | 87,15 | 174,86 | 99,78 | **0,0031** | 350 | 202 | 97 | 97 |
| | 800nm | 139,74 | 72,33 | 165,97 | 84,89 | **0,0003** | 350 | 202 | 97 | 97 |
| | 850nm | 142,52 | 79,66 | 169,39 | 93,30 | **0,0002** | 350 | 202 | 97 | 97 |
| | 910nm | 174,64 | 141,85 | 169,19 | 145,02 | 0,4519 | 341 | 200 | 95 | 96 |
| | 930nm | 214,25 | 179,75 | 200,44 | 171,83 | 0,0565 | 333 | 193 | 93 | 93 |
| | 950nm | 145,19 | 132,88 | 157,49 | 143,58 | 0,0892 | 348 | 201 | 97 | 97 |
| | 980nm | 132,13 | 136,93 | 143,20 | 146,89 | 0,1263 | 341 | 199 | 95 | 96 |
| | 1030nm | 148,22 | 140,90 | 147,81 | 142,97 | 0,9548 | 330 | 192 | 92 | 92 |
| | 1080nm | 123,13 | 103,54 | 127,65 | 109,03 | 0,5321 | 345 | 200 | 96 | 96 |
| | 1100nm | 121,20 | 101,62 | 126,79 | 107,33 | 0,4399 | 348 | 202 | 97 | 97 |
| SWL top10% | 700nm | 327,35 | 179,38 | 353,61 | 196,35 | **0,0089** | 350 | 202 | 97 | 97 |
| | 730nm | 286,45 | 152,74 | 305,28 | 166,63 | 0,0605 | 350 | 202 | 97 | 97 |
| | 760nm | 268,21 | 143,17 | 282,63 | 155,69 | 0,1505 | 350 | 202 | 97 | 97 |
| | 800nm | 234,80 | 113,33 | 250,18 | 125,73 | 0,1254 | 350 | 202 | 97 | 97 |
| | 850nm | 225,39 | 110,39 | 240,12 | 123,29 | 0,1422 | 350 | 202 | 97 | 97 |
| | 910nm | 254,73 | 178,36 | 240,74 | 174,63 | 0,1632 | 344 | 200 | 96 | 96 |
| | 930nm | 311,60 | 225,73 | 288,98 | 209,13 | **0,0241** | 335 | 194 | 93 | 93 |
| | 950nm | 232,24 | 177,97 | 241,23 | 183,70 | 0,3703 | 348 | 201 | 97 | 97 |
| | 980nm | 236,10 | 200,75 | 245,94 | 207,88 | 0,3265 | 345 | 200 | 96 | 96 |
| | 1030nm | 220,96 | 178,23 | 214,43 | 173,67 | 0,5150 | 332 | 193 | 92 | 93 |
| | 1080nm | 176,08 | 127,77 | 176,60 | 127,28 | 0,9585 | 349 | 201 | 97 | 97 |
| | 1100nm | 179,10 | 128,26 | 180,58 | 128,54 | 0,8830 | 349 | 202 | 97 | 97 |
| Spectral Unmixing | Collagen_mean | 1332,39 | 800,71 | 1523,86 | 928,80 | 0,1073 | 350 | 202 | 97 | 97 |
| | Lipids_mean | 1058,88 | 1351,00 | 488,21 | 597,19 | **<0,0001** | 287 | 162 | 80 | 78 |
| | Collagen_top10% | 2099,04 | 1078,56 | 2213,82 | 1172,20 | >0,9999 | 350 | 202 | 97 | 97 |
| | Lipids_top10% | 2356,27 | 2091,45 | 1613,41 | 1326,44 | **<0,0001** | 288 | 163 | 80 | 78 |
| | Hb_mean | 0,0788 | 0,0421 | 0,0887 | 0,0485 | 0,5803 | 347 | 202 | 96 | 97 |
| | Hb_top10% | 0,1654 | 0,0936 | 0,1769 | 0,1013 | 0,2846 | 350 | 202 | 97 | 97 |
| | HbO2_mean | 0,0909 | 0,0507 | 0,1094 | 0,0598 | **0,0063** | 348 | 202 | 97 | 97 |
| | HbO2_top10% | 0,1372 | 0,0636 | 0,1461 | 0,0712 | 0,8363 | 350 | 202 | 97 | 97 |
| | HbT_mean | 0,1689 | 0,0913 | 0,1978 | 0,1068 | **<0,0001** | 349 | 202 | 97 | 97 |
| | HbT_top10% | 0,2756 | 0,1371 | 0,2932 | 0,1519 | **0,0108** | 350 | 202 | 97 | 97 |
| | mSO2_mean | 0,5436 | 0,0706 | 0,5586 | 0,0581 | 0,0514 | 350 | 202 | 97 | 97 |
| | mSO2_top10% | 0,7804 | 0,1133 | 0,7709 | 0,1095 | 0,6543 | 269 | 174 | 75 | 84 |

**Supplementary Table 1. MSOT parameters in FSHD and control muscles.**
Single-wavelength and spectrally unmixed MSOT parameters are compared between muscles from patients with FSHD and healthy controls. Values are reported as mean ± standard deviation. P values indicate between-group comparisons. The number and percentage of analysable measurements are reported for each parameter. Significant P values are highlighted in bold.
