## Supplementary Table 2 for "Optoacoustic molecular signatures of muscle involvement in facioscapulohumeral dystrophy compared to conventional imaging"

| | | T1 = 0 | | T1 = 2-3 | | T1 = 3-4 | | T1 = 0 | T1 = 2-3 | T1 = 3-4 | *P-value* | | |
|---|---|---|---|---|---|---|---|---|---|---|---|---|---|
| | | *Mean* | *SD* | *Mean* | *SD* | *Mean* | *SD* | *N* | *N* | *N* | **T1 = 0 vs. T1 = 1-2** | **T1 = 0 vs. T1 = 3-4** | **T1 = 1-2 vs. T1 = 3-4** |
| SWL mean | 700nm | 175,27 | 93,80 | 128,33 | 72,94 | 131,92 | 69,69 | 123 | 48 | 118 | 0,122 | **0,028** | >0,9999 |
| | 730nm | 162,69 | 86,87 | 119,02 | 68,48 | 124,54 | 64,67 | 124 | 48 | 118 | 0,1842 | 0,0756 | >0,9999 |
| | 760nm | 163,60 | 93,69 | 118,09 | 75,80 | 129,38 | 72,27 | 124 | 48 | 118 | 0,146 | 0,1511 | >0,9999 |
| | 800nm | 154,96 | 79,35 | 110,27 | 61,20 | 111,16 | 52,29 | 124 | 48 | 118 | 0,1622 | **0,0251** | >0,9999 |
| | 850nm | 157,72 | 87,03 | 109,87 | 66,77 | 112,68 | 58,44 | 124 | 48 | 118 | 0,1074 | **0,0193** | >0,9999 |
| | 910nm | 166,63 | 135,66 | 115,51 | 120,68 | 174,51 | 151,55 | 121 | 47 | 116 | 0,0741 | >0,9999 | **0,0247** |
| | 930nm | 201,13 | 160,03 | 136,61 | 151,66 | 227,36 | 203,16 | 115 | 47 | 113 | **0,0103** | 0,5746 | **<0,0001** |
| | 950nm | 147,49 | 133,35 | 91,70 | 98,11 | 133,43 | 132,98 | 123 | 48 | 117 | **0,0349** | >0,9999 | 0,2434 |
| | 980nm | 132,62 | 135,08 | 79,50 | 92,08 | 120,11 | 139,75 | 121 | 47 | 117 | 0,056 | >0,9999 | 0,2875 |
| | 1030nm | 141,37 | 133,28 | 94,26 | 106,37 | 149,07 | 153,69 | 116 | 44 | 113 | 0,1512 | >0,9999 | 0,0569 |
| | 1080nm | 121,16 | 102,57 | 80,18 | 83,83 | 117,99 | 108,29 | 123 | 48 | 118 | 0,2566 | >0,9999 | 0,379 |
| | 1100nm | 121,15 | 101,18 | 81,24 | 81,09 | 116,03 | 106,19 | 123 | 48 | 118 | 0,2912 | >0,9999 | 0,5244 |
| SWL top10% | 700nm | 359,96 | 202,34 | 269,62 | 153,99 | 293,55 | 167,99 | 124 | 48 | 118 | **0,0083** | **0,0113** | >0,9999 |
| | 730nm | 314,08 | 172,47 | 237,25 | 133,89 | 257,95 | 141,00 | 124 | 48 | 118 | **0,0391** | 0,0517 | >0,9999 |
| | 760nm | 290,39 | 159,98 | 218,43 | 128,81 | 242,87 | 131,98 | 124 | 48 | 118 | 0,065 | 0,1569 | >0,9999 |
| | 800nm | 258,29 | 128,34 | 195,27 | 102,05 | 207,26 | 97,52 | 124 | 48 | 118 | 0,1538 | 0,1014 | >0,9999 |
| | 850nm | 244,21 | 122,75 | 185,09 | 101,60 | 199,20 | 95,40 | 124 | 48 | 118 | 0,2178 | 0,2108 | >0,9999 |
| | 910nm | 247,15 | 171,29 | 187,84 | 167,28 | 261,45 | 194,83 | 121 | 47 | 116 | 0,2266 | >0,9999 | 0,0623 |
| | 930nm | 298,91 | 201,39 | 221,54 | 211,21 | 331,52 | 258,38 | 115 | 47 | 113 | **0,0429** | 0,8298 | **0,0008** |
| | 950nm | 236,82 | 180,56 | 164,94 | 152,52 | 223,07 | 187,18 | 123 | 48 | 117 | 0,0661 | >0,9999 | 0,2471 |
| | 980nm | 244,13 | 201,07 | 165,35 | 164,52 | 224,91 | 216,63 | 121 | 47 | 117 | **0,0348** | >0,9999 | 0,2272 |
| | 1030nm | 214,04 | 168,43 | 161,84 | 157,07 | 225,66 | 200,96 | 116 | 44 | 113 | 0,4555 | >0,9999 | 0,1838 |
| | 1080nm | 172,77 | 125,81 | 130,41 | 117,85 | 177,18 | 136,68 | 123 | 48 | 118 | 0,8041 | >0,9999 | 0,6003 |
| | 1100nm | 177,42 | 126,87 | 133,60 | 116,57 | 178,87 | 138,32 | 123 | 48 | 118 | 0,7267 | >0,9999 | 0,6682 |
| Spectral Unmixing | Collagen_mean | 1432,55 | 871,08 | 1037,30 | 701,87 | 1156,07 | 710,86 | 124 | 48 | 118 | 0,5159 | >0,9999 | >0,9999 |
| | Lipids_mean | 596,15 | 596,66 | 701,50 | 988,67 | 1681,48 | 1741,07 | 96 | 16 | 101 | >0,9999 | **<0,0001** | **0,0062** |
| | Collagen_top10% | 2205,82 | 1160,55 | 1748,44 | 1011,77 | 1949,12 | 1005,17 | 124 | 48 | 118 | 0,0512 | >0,9999 | 0,2749 |
| | Lipids_top10% | 1854,12 | 1331,65 | 1886,51 | 1859,03 | 3127,60 | 2630,02 | 96 | 34 | 101 | >0,9999 | **<0,0001** | **<0,0001** |
| | Hb_mean | 0,0833 | 0,0452 | 0,0618 | 0,0363 | 0,0659 | 0,0364 | 124 | 48 | 118 | 0,4207 | 0,3413 | >0,9999 |
| | Hb_top10% | 0,1807 | 0,1053 | 0,1362 | 0,0806 | 0,1509 | 0,0901 | 124 | 48 | 118 | 0,1344 | 0,0943 | >0,9999 |
| | HbO2_mean | 0,1015 | 0,0564 | 0,0690 | 0,0431 | 0,0703 | 0,0361 | 124 | 48 | 118 | 0,0775 | **0,0141** | >0,9999 |
| | HbO2_top10% | 0,1455 | 0,0686 | 0,1130 | 0,0593 | 0,1241 | 0,0577 | 124 | 48 | 118 | **0,0074** | **0,0004** | >0,9999 |
| | HbT_mean | 0,1849 | 0,1001 | 0,1308 | 0,0779 | 0,1362 | 0,0696 | 124 | 48 | 118 | **0,0006** | **<0,0001** | >0,9999 |
| | HbT_top10% | 0,3007 | 0,1538 | 0,2269 | 0,1243 | 0,2446 | 0,1198 | 124 | 48 | 118 | **<0,0001** | **<0,0001** | >0,9999 |
| | mSO2_mean | 0,5647 | 0,0668 | 0,5332 | 0,0628 | 0,5205 | 0,0737 | 124 | 48 | 118 | 0,0919 | **0,0002** | >0,9999 |
| | mSO2_top10% | 0,7751 | 0,1134 | 0,7833 | 0,1130 | 0,8101 | 0,1143 | 95 | 32 | 83 | **0,0099** | **<0,0001** | 0,7166 |

**Supplementary Table 2. MSOT parameters according to MRI T1-weighted fatty replacement.**
Single-wavelength and spectrally unmixed MSOT parameters are shown after stratification of FSHD muscles by MRI T1 score. Muscles were grouped as T1 = 0, T1 = 1–2 and T1 = 3–4. Values are reported as mean ± standard deviation. P values indicate post hoc comparisons between MRI-defined fatty replacement categories. Significant P values are highlighted in bold.
