## Supplementary Table 3 for "Optoacoustic molecular signatures of muscle involvement in facioscapulohumeral dystrophy compared to conventional imaging"

| | | STIR+ | | STIR- | | | STIR+ | STIR- |
|---|---|---|---|---|---|---|---|---|
| | | *Mean* | *SD* | *Mean* | *SD* | *P-value* | *N* | *N* |
| SWL mean | 700nm | 174,61 | 83,71 | 144,70 | 83,30 | 0,0915 | 49 | 240 |
| | 730nm | 163,28 | 78,06 | 135,20 | 77,10 | 0,1130 | 49 | 241 |
| | 760nm | 167,15 | 83,81 | 137,06 | 83,93 | 0,0896 | 49 | 241 |
| | 800nm | 146,73 | 68,18 | 126,29 | 69,76 | 0,2486 | 49 | 241 |
| | 850nm | 150,10 | 74,14 | 127,68 | 76,54 | 0,2057 | 49 | 241 |
| | 910nm | 206,13 | 144,20 | 152,29 | 139,13 | **0,0026** | 48 | 236 |
| | 930nm | 258,09 | 188,51 | 189,09 | 176,28 | **0,0001** | 47 | 228 |
| | 950nm | 173,56 | 134,68 | 124,06 | 126,53 | **0,0053** | 49 | 239 |
| | 980nm | 161,71 | 143,45 | 109,80 | 127,90 | **0,0035** | 49 | 236 |
| | 1030nm | 181,01 | 146,33 | 127,81 | 136,29 | **0,0033** | 47 | 226 |
| | 1080nm | 146,41 | 106,83 | 106,25 | 100,94 | **0,0235** | 49 | 240 |
| | 1100nm | 145,60 | 105,37 | 105,66 | 98,89 | **0,0243** | 49 | 240 |
| SWL top10% | 700nm | 378,25 | 170,38 | 306,29 | 185,54 | **0,0051** | 49 | 241 |
| | 730nm | 332,51 | 148,41 | 267,54 | 156,61 | **0,0114** | 49 | 241 |
| | 760nm | 311,69 | 142,27 | 248,46 | 145,25 | **0,0137** | 49 | 241 |
| | 800nm | 266,91 | 113,39 | 219,00 | 114,26 | 0,0619 | 49 | 241 |
| | 850nm | 256,77 | 113,02 | 207,84 | 109,36 | 0,0565 | 49 | 241 |
| | 910nm | 314,96 | 187,15 | 228,58 | 177,62 | **0,0009** | 48 | 236 |
| | 930nm | 388,70 | 240,74 | 280,61 | 224,43 | **<0,0001** | 47 | 228 |
| | 950nm | 295,77 | 193,05 | 203,57 | 173,71 | **0,0003** | 49 | 239 |
| | 980nm | 311,19 | 226,93 | 204,99 | 193,74 | **<0,0001** | 49 | 236 |
| | 1030nm | 286,40 | 195,11 | 194,64 | 175,11 | **0,0005** | 47 | 226 |
| | 1080nm | 222,76 | 139,31 | 156,26 | 125,07 | **0,0096** | 49 | 240 |
| | 1100nm | 229,08 | 142,14 | 158,82 | 125,25 | **0,0062** | 49 | 240 |
| Spectral Unmixing | Collagen_mean | 1480,11 | 789,12 | 1208,79 | 790,72 | 0,5356 | 49 | 241 |
| | Lipids_mean | 1374,92 | 1513,02 | 1034,57 | 1361,05 | >0,9999 | 44 | 187 |
| | Collagen_top10% | 2593,63 | 1801,06 | 1709,08 | 1553,13 | **0,0012** | 49 | 241 |
| | Lipids_top10% | 3332,25 | 2512,03 | 2351,42 | 2088,27 | **0,0004** | 46 | 214 |
| | Hb_mean | 0,0859 | 0,0415 | 0,0700 | 0,0409 | >0,9999 | 49 | 241 |
| | Hb_top10% | 0,1932 | 0,0889 | 0,1547 | 0,0972 | **0,0378** | 49 | 241 |
| | HbO2_mean | 0,0933 | 0,0466 | 0,0815 | 0,0497 | >0,9999 | 49 | 241 |
| | HbO2_top10% | 0,1535 | 0,0666 | 0,1269 | 0,0625 | 0,4139 | 49 | 241 |
| | HbT_mean | 0,1792 | 0,0868 | 0,1514 | 0,0883 | 0,3361 | 49 | 241 |
| | HbT_top10% | 0,3153 | 0,1378 | 0,2556 | 0,1375 | **0,0013** | 49 | 241 |
| | mSO2_mean | 0,5278 | 0,0492 | 0,5442 | 0,0754 | >0,9999 | 49 | 241 |
| | mSO2_top10% | 0,8226 | 0,1058 | 0,7841 | 0,1151 | 0,1587 | 33 | 177 |

**Supplementary Table 3. MSOT parameters according to STIR status.**
Single-wavelength and spectrally unmixed MSOT parameters are compared between STIR-positive and STIR-negative FSHD muscles. Values are reported as mean ± standard deviation. P values indicate between-group comparisons according to STIR status. Significant P values are highlighted in bold.
