## Supplementary Table 4 for "Optoacoustic molecular signatures of muscle involvement in facioscapulohumeral dystrophy compared to conventional imaging"

| | | **Heckmatt = 1** | | **Heckmatt = 2-3** | | **Heckmatt = 4** | | **Heckmatt = 1** | **Heckmatt = 2-3** | **Heckmatt = 4** | *P-value* | | |
|---|---|---|---|---|---|---|---|---|---|---|---|---|---|
| | | *Mean* | *SD* | *Mean* | *SD* | *Mean* | *SD* | *N* | *N* | *N* | **Heckmatt 1 vs 4** | **Heckmatt 1 vs 2-3** | **Heckmatt 2-3 vs 4** |
| SWL mean | 700nm | 188,80 | 94,45 | 127,85 | 71,00 | 140,49 | 73,29 | 165 | 28 | 155 | **0,0017** | 0,0728 | >0,9999 |
| | 730nm | 174,18 | 86,60 | 116,81 | 65,30 | 132,08 | 67,99 | 165 | 28 | 155 | **0,0093** | 0,1093 | >0,9999 |
| | 760nm | 177,23 | 93,78 | 112,31 | 68,63 | 136,68 | 75,39 | 165 | 28 | 155 | 0,0138 | **0,0453** | >0,9999 |
| | 800nm | 164,55 | 79,05 | 108,29 | 59,89 | 119,77 | 57,06 | 165 | 28 | 155 | **0,0046** | 0,1236 | >0,9999 |
| | 850nm | 169,42 | 87,23 | 103,89 | 62,87 | 121,89 | 63,43 | 165 | 28 | 155 | **0,0021** | **0,0421** | >0,9999 |
| | 910nm | 185,45 | 140,42 | 86,04 | 84,90 | 177,92 | 147,12 | 164 | 27 | 151 | >0,9999 | **0,0003** | **0,0013** |
| | 930nm | 220,15 | 169,81 | 96,10 | 98,61 | 230,25 | 194,34 | 160 | 27 | 146 | >0,9999 | **<0,0001** | **<0,0001** |
| | 950nm | 168,27 | 139,81 | 72,24 | 76,90 | 135,99 | 127,72 | 164 | 28 | 154 | 0,0933 | **0,0005** | 0,0544 |
| | 980nm | 153,01 | 145,04 | 58,66 | 71,85 | 121,87 | 131,83 | 163 | 28 | 152 | 0,1211 | **0,0006** | 0,0584 |
| | 1030nm | 158,96 | 140,90 | 60,42 | 73,24 | 152,28 | 145,49 | 159 | 27 | 145 | >0,9999 | **0,0004** | **0,0014** |
| | 1080nm | 135,80 | 105,17 | 62,55 | 63,04 | 119,37 | 104,45 | 164 | 28 | 155 | >0,9999 | **0,0156** | 0,1198 |
| | 1100nm | 135,85 | 103,31 | 63,16 | 61,11 | 117,21 | 101,99 | 164 | 28 | 155 | 0,9693 | **0,0168** | 0,1611 |
| SWL top10% | 700nm | 366,86 | 183,93 | 238,76 | 134,05 | 301,98 | 168,40 | 165 | 28 | 155 | **0,0024** | **0,0008** | 0,3617 |
| | 730nm | 319,73 | 156,97 | 210,86 | 116,75 | 265,26 | 142,48 | 165 | 28 | 155 | **0,0178** | **0,0069** | 0,6361 |
| | 760nm | 298,64 | 146,67 | 192,94 | 111,03 | 250,20 | 134,15 | 165 | 28 | 155 | **0,0495** | **0,0096** | 0,5329 |
| | 800nm | 262,85 | 118,75 | 178,56 | 92,77 | 215,65 | 100,56 | 165 | 28 | 155 | 0,0603 | 0,0712 | >0,9999 |
| | 850nm | 252,38 | 115,51 | 166,93 | 89,22 | 208,17 | 99,10 | 165 | 28 | 155 | 0,0954 | 0,0645 | >0,9999 |
| | 910nm | 262,93 | 168,61 | 144,48 | 112,45 | 268,39 | 191,01 | 164 | 27 | 151 | >0,9999 | **0,003** | **0,0018** |
| | 930nm | 313,77 | 204,89 | 163,64 | 132,39 | 340,63 | 249,26 | 160 | 27 | 146 | 0,9125 | **<0,0001** | **<0,0001** |
| | 950nm | 256,33 | 176,19 | 134,72 | 116,71 | 226,98 | 182,94 | 164 | 28 | 154 | 0,6629 | **0,0017** | **0,0369** |
| | 980nm | 263,31 | 197,84 | 129,88 | 131,20 | 229,80 | 207,97 | 163 | 28 | 152 | 0,4188 | **0,0004** | **0,0182** |
| | 1030nm | 228,67 | 168,64 | 109,34 | 102,03 | 234,87 | 192,72 | 159 | 27 | 145 | >0,9999 | **0,0028** | **0,0016** |
| | 1080nm | 187,35 | 123,49 | 101,60 | 81,54 | 179,93 | 134,67 | 164 | 28 | 155 | >0,9999 | 0,063 | 0,1196 |
| | 1100nm | 192,01 | 123,92 | 106,81 | 82,87 | 180,77 | 135,39 | 164 | 28 | 155 | >0,9999 | 0,0661 | 0,1678 |
| Spectral Unmixing | Collagen_mean | 1516,15 | 863,00 | 937,40 | 615,82 | 1222,09 | 711,63 | 165 | 28 | 155 | 0,2325 | 0,156 | >0,9999 |
| | Lipids_mean | 665,94 | 828,55 | 222,55 | 357,69 | 1596,44 | 1656,33 | 139 | 19 | 130 | **<0,0001** | 0,9243 | **<0,0001** |
| | Collagen_top10% | 2290,40 | 1136,76 | 1510,42 | 797,13 | 2015,94 | 1005,91 | 165 | 28 | 155 | 0,3222 | **0,0162** | 0,3175 |
| | Lipids_top10% | 1884,35 | 1516,73 | 932,03 | 924,55 | 3066,61 | 2476,36 | 139 | 19 | 130 | **<0,0001** | **0,0133** | **<0,0001** |
| | Hb_mean | 0,0896 | 0,0454 | 0,0614 | 0,0343 | 0,0692 | 0,0375 | 165 | 28 | 155 | 0,2358 | 0,709 | >0,9999 |
| | Hb_top10% | 0,1839 | 0,0952 | 0,1194 | 0,0687 | 0,1542 | 0,0899 | 165 | 28 | 155 | **0,0157** | **0,0021** | 0,3281 |
| | HbO2_mean | 0,1082 | 0,0560 | 0,0640 | 0,0396 | 0,0766 | 0,0398 | 165 | 28 | 155 | **0,0081** | 0,0851 | >0,9999 |
| | HbO2_top10% | 0,1515 | 0,0659 | 0,1013 | 0,0497 | 0,1294 | 0,0591 | 165 | 28 | 155 | 0,1511 | **0,0325** | 0,7276 |
| | HbT_mean | 0,1978 | 0,0996 | 0,1104 | 0,0602 | 0,1458 | 0,0746 | 165 | 25 | 155 | **<0,0001** | **<0,0001** | 0,3758 |
| | HbT_top10% | 0,3086 | 0,1428 | 0,2047 | 0,1102 | 0,2544 | 0,1235 | 165 | 28 | 155 | **<0,0001** | **<0,0001** | **0,0369** |
| | mSO2_mean | 0,5598 | 0,0664 | 0,5101 | 0,0607 | 0,5320 | 0,0727 | 165 | 25 | 155 | **0,0294** | 0,0524 | >0,9999 |
| | mSO2_top10% | 0,7681 | 0,1125 | 0,7581 | 0,1264 | 0,8011 | 0,1054 | 133 | 22 | 111 | **0,022** | >0,9999 | 0,2198 |

**Supplementary Table 4. MSOT parameters according to RUCT Heckmatt grade.**
Single-wavelength and spectrally unmixed MSOT parameters are shown after stratification by RUCT-based Heckmatt grading. Muscles were grouped as Heckmatt grade 1, grades 2–3 and grade 4. Values are reported as mean ± standard deviation. P values indicate post hoc comparisons between Heckmatt categories. Significant P values are highlighted in bold.
