## Supplementary Table 5 for "Optoacoustic molecular signatures of muscle involvement in facioscapulohumeral dystrophy compared to conventional imaging"

| Dynamometry | | | | |
|---|---|---|---|---|
| | | *Spearman r correlation coefficient* | *95% confidence interval* | *P-value* |
| SWL mean | 700nm | 0,2394 | 0,1015 to 0,3683 | **0,0006** |
| | 730nm | 0,2126 | 0,07342 to 0,3436 | **0,0023** |
| | 760nm | 0,1658 | 0,02499 to 0,3001 | **0,0178** |
| | 800nm | 0,2444 | 0,1067 to 0,3729 | **0,0004** |
| | 850nm | 0,2091 | 0,06983 to 0,3405 | **0,0027** |
| | 910nm | -0,07469 | -0,2155 to 0,06920 | 0,2944 |
| | 930nm | -0,1065 | -0,2470 to 0,03830 | 0,1373 |
| | 950nm | -0,003478 | -0,1451 to 0,1383 | 0,9607 |
| | 980nm | -0,0248 | -0,1666 to 0,1180 | 0,7267 |
| | 1030nm | -0,09636 | -0,2369 to 0,04818 | 0,178 |
| | 1080nm | -0,04215 | -0,1828 to 0,1002 | 0,5505 |
| | 1100nm | -0,03802 | -0,1788 to 0,1043 | 0,5902 |
| SWL top10% | 700nm | 0,1413 | -0,0001288 to 0,2771 | **0,0439** |
| | 730nm | 0,1242 | -0,01752 to 0,2610 | 0,0768 |
| | 760nm | 0,1091 | -0,03281 to 0,2467 | 0,1204 |
| | 800nm | 0,1367 | -0,004727 to 0,2728 | 0,0511 |
| | 850nm | 0,1067 | -0,03520 to 0,2444 | 0,1287 |
| | 910nm | -0,129 | -0,2672 to 0,01444 | 0,0695 |
| | 930nm | -0,1533 | -0,2911 to -0,009267 | **0,0319** |
| | 950nm | -0,0351 | -0,1760 to 0,1072 | 0,6191 |
| | 980nm | -0,0488 | -0,1899 to 0,09428 | 0,4915 |
| | 1030nm | -0,139 | -0,2773 to 0,004984 | 0,0514 |
| | 1080nm | -0,09356 | -0,2322 to 0,04882 | 0,1843 |
| | 1100nm | -0,08472 | -0,2238 to 0,05770 | 0,2294 |
| Spectral Unmixing | Collagen_mean | -0,04308 | -0,1840 to 0,09961 | 0,5427 |
| | Lipids_mean | -0,264 | -0,3939 to -0,1237 | **0,0002** |
| | Collagen_top10% | -0,06063 | -0,2010 to 0,08215 | 0,3913 |
| | Lipids_top10% | -0,2738 | -0,4028 to -0,1341 | **0,0001** |
| | Hb_mean | 0,2017 | 0,06170 to 0,3338 | **0,0039** |
| | Hb_top10% | 0,1091 | -0,03313 to 0,2470 | 0,1212 |
| | HbO2_mean | 0,1673 | 0,02617 to 0,3018 | **0,0171** |
| | HbO2_top10% | 0,02115 | -0,1209 to 0,1624 | 0,7645 |
| | HbT_mean | 0,2001 | 0,06011 to 0,3324 | **0,0042** |
| | HbT_top10% | 0,1098 | -0,03239 to 0,2477 | 0,1187 |
| | mSO2_mean | 0,04226 | -0,1001 to 0,1829 | 0,5494 |
| | mSO2_top10% | -0,2414 | -0,3833 to -0,08850 | **0,0017** |

| CSS | | | | |
|---|---|---|---|---|
| | | *Spearman r correlation coefficient* | *95% confidence interval* | *P-value* |
| Spectral Unmixing | Collagen_mean | -0,50190 | 0,2325 to 0,9189 | 0,05874 |
| | Lipids_mean | 0,13639 | -0,6643 to 0,4966 | 0,62528 |
| | Collagen_top10% | -0,51099 | 0,3351 to 0,9346 | 0,05380 |
| | Lipids_top10% | 0,00727 | -0,5333 to 0,6355 | 0,98176 |
| | Hb_mean | -0,49463 | 0,1589 to 0,9061 | 0,06293 |
| | Hb_top10% | -0,54373 | 0,2465 to 0,9212 | **0,03858** |
| | HbO2_mean | -0,60192 | 0,2973 to 0,9291 | **0,01986** |
| | HbO2_top10% | -0,47281 | 0,2187 to 0,9166 | 0,07684 |
| | HbT_mean | -0,60374 | 0,2325 to 0,9189 | **0,01942** |
| | HbT_top10% | -0,54009 | 0,4321 to 0,9476 | **0,04009** |
| | mSO2_mean | -0,05274 | -0,4884 to 0,6703 | 0,85218 |
| | mSO2_top10% | -0,07638 | -0,2642 to 0,7912 | 0,78607 |

**Supplementary Table 5. Correlations between MSOT-derived parameters, muscle strength and clinical severity.**
Spearman correlation analyses between MSOT-derived single-wavelength and spectrally unmixed parameters, hand-held dynamometry and FSHD Clinical Severity Scale score are reported. Dynamometry correlations were performed at the muscle level using strength values from the corresponding muscle. Clinical Severity Scale correlations were performed at the patient level using summary MSOT measures averaged across assessed muscles. Spearman r coefficients, 95% confidence intervals and P values are shown. Significant P values are highlighted in bold.
